# Spinal Manipulation and Other Conservative Treatments for Acute and Chronic Low Back Pain Compared with Natural History and Sham: A Bayesian Network Meta-Analysis

**DOI:** 10.64898/2026.09.16.26359287

**Authors:** James Michael Menke, Aurelio José Figueredo, Mateo Peñaherrera-Aguirre

## Abstract

**Background:** Patients with low back pain (LBP) often improve during treatment. But some of that improvement would have happened anyway (natural recovery). Some comes from the experience of receiving care (contextual response). Only the rest is due to the treatment itself. We evaluated spinal manipulative therapy (SMT) and common alternatives in two ways: against each other, and against an independent estimate of how LBP improves without treatment (natural history).

**Methods:** We conducted a Bayesian hierarchical network meta-analysis (NMA) of 133 retained analytic units. Acute and chronic LBP were analyzed separately. A companion paper (Paper I) estimated natural history for each phase from separate prognosis studies. We imported those estimates unchanged. No treatment-trial data were used to build them. They served as an external “what would have happened anyway” comparison. The NMA compared SMT, exercise, medical care, physical modalities, self-care, and sham. We kept two comparisons separate: treatment minus natural history, and treatment minus sham. Uncertainty from both analyses was carried through to the final comparisons.

**Results:** In acute LBP, natural history predicted that 75.2% of patients would recover by 26 weeks. SMT recovery was 63.2%. That is 12.0 fewer recoveries per 100 than natural history (95% interval −23.2 to −0.7) and 3.1 more per 100 than sham (−6.3 to +12.8). In chronic LBP, where recovery means a favorable course, natural-history recovery at 54 weeks was 18.1%. SMT recovery was 31.8%. That is 13.8 more recoveries per 100 than natural history (+4.3 to +22.6) and 13.4 more than sham (+4.3 to +21.9). Exercise was similar (+12.5 per 100 versus sham; +5.5 to +20.4). Chronic sham and natural history were nearly identical (+0.4 per 100; −4.2 to +5.7).

**Conclusions:** Whether an LBP treatment looks effective depends on its comparator. Our linked design keeps the prognosis evidence and the treatment evidence apart until the final comparison. In acute LBP, recovery rates were high, and no treatment added a detectable benefit beyond sham or beyond expected natural recovery. In chronic LBP, SMT and exercise showed some benefit beyond natural history and sham. Even so, only about one in three chronic patients was modeled to recover with either treatment.

## Introduction

Low back pain (LBP) is a leading cause of disability and lost work worldwide [1]. In the United States, low back and neck pain accounted for more health-care spending than any other condition, an estimated $134.5 billion in 2016 [2]. Decades of randomized trials and many systematic reviews have failed to settle which conservative treatments work best. Spinal manipulative therapy (SMT), exercise, medical care, physical modalities, education, and self-care have each been reported superior in some of five decades of studies. Yet the differences among them are inconsistent from one trial or review to the next[1–6].

One reason may be simple. Getting better during treatment is not the same as getting better because of treatment. Changes in pain can have several sources. The condition may be improving on its own. Patients often enroll in trials during a flare, and pain tends to drift back toward its usual level afterward (regression to the mean). Patients may also respond to attention, expectation, reassurance, and simply taking part in care. Only the remainder is caused by the treatment itself. Randomized trials can estimate differences between treatment groups. They cannot, by themselves, tell us how much of the improvement within each group would have happened without the treatment [7–9].

This matters greatly in LBP because the natural course depends on how long the pain has lasted. Acute episodes often improve a great deal on their own. Chronic LBP improves more slowly and less completely. So, a treatment that looks impressive in acute LBP may add little to recovery that would have happened anyway. The same observed improvement in chronic LBP may mean much more, because it departs further from the expected course. If we pool or interpret treatment effects without modeling this background recovery, we can miss important differences between acute and chronic LBP [10–37].

A second problem is the choice of comparator. In this analysis, “sham” means a placebo procedure or a treatment known to be ineffective. Patients who receive sham still improve. That improvement has two parts: the natural course of LBP, and the response to being treated in a clinical or research setting. The NMA compared all treatments with one another. Separately, the independent natural-history estimate showed how much improvement to expect without the treatments being compared. Subtracting natural history from sham (sham minus NH) gave us an estimate of placebo responsiveness.

A third problem is that the trials differ a great deal (heterogeneity). They enroll different patients, follow them for different lengths of time, and deliver treatment at different intensities. Providers come from different professions. Study quality and sham designs vary. Factors such as expectation, confidence, trust, and the patient–provider relationship may matter, but they are rarely measured or reported. These differences can distort both direct and indirect comparisons. They can also undermine two assumptions that an NMA depends on: transitivity and exchangeability (explained in the Methods). Differences between studies must therefore be modeled directly, not assumed to be differences between treatments.

This study uses a Bayesian hierarchical model and analyzes acute and chronic LBP separately. The natural-history (NH) trajectories come from a companion meta-analysis (Paper I). Their uncertainty is carried forward into the treatment analysis.

The guiding question is simple: compared with what? Getting better, getting better while being treated, getting better more than expected, and getting better more than with sham are related ideas. But they are not the same causal claim. The analysis therefore has two separate Bayesian parts. Prognosis studies produce the natural-history estimates. Treatment trials produce the treatment estimates. The two meet only at the end, when we compare their posterior distributions directly. Natural history is not added to the network as an extra control arm. Treatment minus natural history and treatment minus sham are different comparisons, and they cannot simply be added together. Paper I estimates the natural-history posteriors [38]. This paper applies them to SMT and its comparators.

## Objectives

Our objectives were to: (1) compare SMT with other conservative treatments in separate acute and chronic Bayesian networks; (2) estimate treatment and sham trajectories without using the prognosis evidence; (3) import the natural-history estimates from Paper I and compare treatments with them, but only when both measure the same outcome in the same way; (4) estimate treatment minus sham and sham minus natural history as separate comparisons; and (5) test how robust the results are to study quality, heterogeneity, transitivity, exchangeability, the definition of recovery, and direct versus indirect evidence.

To our knowledge, this is the first SMT analysis to estimate natural recovery independently and carry its uncertainty into a comparison of treatments.

## Methods

### Study design and analytic framework

We combined comparative studies of conservative treatments for low back pain (LBP) in a Bayesian hierarchical model. Randomized and controlled clinical trials were the main source of evidence. We also kept some prospective observational cohorts when they added useful information about how pain changes over time or how much studies differ. We did not treat these cohorts as if they had been randomized, so we did not assume they were free of confounding.

The analysis was built to separate three parts of observed improvement. The first is improvement expected from the natural course of LBP. The second is improvement from the general experience of receiving care. The third is improvement due to the specific treatment. Acute and chronic LBP were analyzed separately. Their natural courses differ a great deal, and pooling them would mix patients who are not truly comparable.

### Dual Bayesian analytic logic

This paper is the second of two linked Bayesian analyses. Paper I combines separate prognosis studies to estimate natural history for each phase of LBP. This paper does not re-estimate those models. Instead, it builds its own Bayesian treatment networks for each phase. It brings in the Paper I results only at the final, interpretive step.

We kept the two evidence streams apart. Treatment trials do not update the natural-history estimate. Sham arms are not relabeled as natural history. Prognosis cohorts are not added to the network as stand-in control arms. Results from the two analyses are combined only when they describe the same clinical outcome. Treatment versus sham is a separate comparison made within the network.

This separation matters most for chronic LBP. Chronic pain comes and goes. So “meaningful improvement,” “time spent doing well,” “pain-free,” “remission,” “persistence,” and “recurrence” are different outcomes. They cannot be swapped for one another. We therefore report treatment minus natural history only when the chronic natural history estimate and the treatment estimate measure the same outcome. Otherwise, they must be linked by an explicit and defensible measurement model. We did this by fitting the chronic treatment model with Paper I’s chronic natural-history posterior, in its reference outcome, as the starting line (see Outcomes).

The final evidence set had 133 analytic units from 131 parent trials. (An analytic unit is a study, or a separable part of a study, that contributes data.) Of these, 59 contributed acute data and 74 contributed chronic data. Three units contributed to both. That gives 130 unique units assigned to a phase. Kinalski [39] had mixed or unknown phase and was not assigned to either analysis.

Sturion [40] was kept to document its design before crossover, but it gave no separate treatment estimate we could use. Licciardone [41] reported a single cross-sectional pain score at registry enrollment, with no follow-up, so it gave no longitudinal estimate and was kept in the inventory only. Each observation was identified by study, treatment category (the T6 variable), and week of follow-up. Each study was assigned to the acute or chronic phase. Acute observations were modeled through 26 weeks and chronic observations through 54 weeks. Table 1 summarizes treatments by phase. Table 2 specifies the Bayesian model. Table 3 accounts for every unit in the evidence base.

**Table 1.** Treatment characteristics of the comparative evidence base. Phase-specific treatment categories are shown with source-arm counts, initial/earliest reported sample size, and PEDro quality scores.

| Phase | Treatment class | Studies /<br>analytic<br>units | Treatment<br>arms | Initial / earliest<br>reported N | PEDro mean (SD) |
| --- | --- | --- | --- | --- | --- |
| **ACUTE** |  |  |  |  |  |
|  | SMT | 59 | 68 | 4,542 | 4.86 (2.14) |
|  | Exercise | 12 | 14 | 764 | 4.33 (1.67) |
|  | Medical | 20 | 22 | 1,538 | 5.26 (2.51) |
|  | Modalities | 23 | 25 | 1,612 | 3.78 (1.81) |
|  | Self-care | 10 | 11 | 922 | 4.60 (2.59) |
|  | Sham | 13 | 15 | 874 | 4.83 (2.55) |
| **CHRONIC** |  |  |  |  |  |
|  | SMT | 70 | 74 | 4,785 | 5.89 (1.86) |
|  | Exercise | 27 | 29 | 1,581 | 5.85 (1.96) |
|  | Medical | 18 | 19 | 1,282 | 5.78 (1.90) |
|  | Modalities | 21 | 22 | 1,104 | 5.76 (1.92) |
|  | Self-care | 4 | 5 | 247 | 5.75 (3.30) |
|  | Sham | 28 | 31 | 2,052 | 6.09 (1.88) |
| **NATURAL HISTORY** |  |  |  |  |  |
|  | Acute Natural History | 8 | — | — | — |
|  | Chronic Natural History | 12 | — | — | — |

The retained evidence base comprised 131 parent trials represented by 133 analytic units; inventory reconciliation is reported separately in Table 3.
Treatment arms are descriptive source arms only. Each analytic observation is one study x treatment x follow-up week.
No genuine week-0 observations were available in the quantitative treatment data; sample size is therefore reported as Initial / earliest reported N rather than as baseline N.
Natural History source evidence comprises 8 acute studies / 26 observations and 12 chronic cohorts / 15 observations (12-cohort chronic model, all follow-up times). Sham is the randomized sham or therapeutically inactive comparator category; Natural History is not a randomized treatment arm and therefore has no treatment-arm count or PEDro summary.

**Table 2.** Hierarchical Bayesian model specification.

| Element | Specification |
| --- | --- |
| **OVERVIEW** |  |
| **Model structure** | Bayesian hierarchical longitudinal treatment model with an externally estimated Bayesian Natural History trajectory, hierarchical treatment deviations, SMT-provider deviations, study random intercepts, and non-centered treatment-by-study random effects. Acute and chronic low back pain are modeled separately. |
| **Primary model input** | Acute: 220 study x treatment x week observations from 46 studies ($\leq 26$ weeks). Chronic: 264 observations from 54 studies ($\leq 54$ weeks). These are model-input counts after time-domain and model-validity filtering, not baseline participant counts. |
| **Analytic observation** | Study x treatment x follow-up week. Multiple source rows mapping to one key are inverse-variance aggregated before Bayesian treatment modeling; sample sizes are summed within the key. |
| **Outcome measure** | Hedges g from within-arm change in pain on a pooled-SD scale (2 points on 0-10 scales; 20 points on 0-100 scales); positive values indicate improvement. Values derived by probit conversion are excluded from the primary continuous-outcome model. No winsorization is applied. |
| **MODEL SPECIFICATION** |  |
| **Likelihood** | $y_i \sim \text{Normal}(\mu_i, \text{se}_i^2 + \text{Var\_NH}(t_i) + \sigma_{\text{resid}}^2)$. The Natural History posterior mean is an offset and its pointwise posterior variance is propagated in the observation variance. |
| **Linear predictor** | $\mu_i = \text{NH}(t_i) + \beta_n \cdot \text{centered log}(n_i) + \beta_q \cdot \text{centered PEDro}_i + \text{tx\_effect}[\text{treatment}_i] + l(\text{SMT}_i) \cdot \gamma[\text{provider}_i] + u[\text{study}_i] + v[\text{treatment}_i, \text{study}_i]$. |
| **Natural History - acute** | Natural-course/prognosis evidence only: 8 studies / 26 observations. $g_{\text{NH}}(t) = \beta_0 + \beta_1 [1 - \exp(-\lambda t)]$. Current posterior means: $\beta_0 = 0.247$, $\beta_1 = 1.926$, $\lambda = 0.39965/\text{week}$. Study random intercept plus random trajectory/rate. Trial data do not update the Natural History trajectory. |
| **Natural History - chronic** | Natural-course/prognosis evidence only: 12 cohorts / 15 observations at all reported follow-up times (54 weeks is the reporting horizon, not an eligibility rule). $\text{logit } p(t) = \theta_{54} + \beta \cdot [\log(1+t) - \log(1+54)] + \alpha_{\text{family}} + u[\text{cohort}]$; outcome-family offsets for STRICT and COURSE definitions, DIRECT is the reference family. Posterior means: $\theta_{54} = -1.561$, $\beta = 0.176$, $\alpha_{\text{strict}} = 0.455$, $\alpha_{\text{course}} = 0.368$, $\sigma_u = 0.857$. This posterior is the chronic treatment model's external NH offset, entered on the g scale as $qnorm(p_{\text{NH}}(t)) + \Delta_{\text{CHRONIC}}$ before treatment effects are estimated; chronic treatment probabilities are $p_k = \Phi(qnorm(p_{\text{NH}}) + \text{tx\_effect}_k)$. |
| **Treatment effects** | Six categories: sham, SMT, exercise, medical care, modalities, and self-care. $\text{tx\_effect}[k]$ is the time-invariant additive deviation from Natural History and is modeled hierarchically as $\text{Normal}(0, \sigma_{\text{tx}}^2)$. |
| **SMT provider effects** | Provider effects use the non-centered hierarchy $\gamma[p] = z_{\gamma[p]} \cdot \sigma_{\text{prov}}$ for PT, DC, DO, MD, Bonesetters, and Mixed; non-SMT observations use the neutral provider code. |
| **Study effects** | $u[\text{study}] \sim \text{Normal}(0, \tau^2)$. Treatment-by-study effects use the non-centered hierarchy $v[k, j] = z_{v[k, j]} \cdot \sigma_{\text{tx\_x\_study}}$. |
| **Covariates** | Centered log(sample size) and centered PEDro score; $\beta_n$ and $\beta_q \sim \text{Normal}(0, 1)$. PEDro is a covariate/sensitivity variable, not an exclusion criterion. |
| **Scale priors** | $\sigma_{\text{tx}}$, $\sigma_{\text{prov}}$, $\tau$, $\sigma_{\text{tx\_x\_study}}$, and $\sigma_{\text{resid}}$ use half-t(df=1, scale=0.5) priors. |
| **TECHNICAL DETAILS** |  |
| **Treatment-model MCMC** | JAGS via rjags; 4 chains. Acute: 5,000 adaptation + 35,000 burn-in + 150,000 sampling, thin=4. Chronic: 4,000 adaptation + 35,000 burn-in + 150,000 sampling, thin=4. Fresh-sample retry is triggered if max PSRF exceeds 1.01. |
| **NH-model MCMC** | JAGS via rjags; 4 chains; 20,000 adaptation. Acute: 40,000 burn-in + 400,000 sampling, thin=10, ESS floor 1,500. Chronic (12-cohort model): 200,000 burn-in + 1,200,000 sampling, thin=50, ESS floor 1,000, sampling doubled automatically (up to 4 attempts) until the gate passes. Both require max PSRF < 1.01. |
| **Convergence** | Maximum PSRF: treatment models 1.0007 (acute) and 1.0026 (chronic); Natural History models 1.0034 (acute) and 1.0029 (chronic). All prespecified convergence criteria (PSRF < 1.01) were met. |
| **INTERPRETATION** |  |
| **Recovery translation** | Acute: recovery = pain $\leq 2/10$; threshold Delta = 1.412 from the weighted acute baseline mean; $P(\text{recovery}) = \Phi(g - \text{Delta})$. Chronic: recovery = favorable natural course in the 12-cohort NH reference (DIRECT) family; $P_k = \Phi(\Phi^{-1}(p_{\text{NH}}) + \text{effect}_k)$, with alternative recovery definitions shifted by (1.642 - Delta_alt). Treatment effects are time-invariant, so no additional time-decay factor is applied. |
| **Reference labels** | Reader-facing contrasts are Natural History and sham. Placebo responsiveness is defined separately as sham minus natural history in the manuscript Methods. |
| **CODING / DATA POLICY** |  |
| **Phase encoding** | Each study was assigned to the acute or chronic phase. Primary time domains are acute $\leq 26$ weeks and chronic $\leq 54$ weeks. Rows without a phase assignment are counted in the evidence-base audit but do not enter phase-specific models. |
| **Treatment coding** | Every treatment arm was coded into one of six categories: sham, SMT, exercise, medical care, modalities, or self-care. Sham includes randomized sham or therapeutically inactive comparators. |
| **Missingness / exclusions** | No imputation or earliest-follow-up substitution is used. Structurally non-longitudinal records or rows lacking a valid study, treatment, week, and sample size are counted in the audit but do not enter longitudinal estimation. For the crossover trial of Sturion 2020, only pre-crossover pain data were eligible. |
| **Sensitivity / diagnostics** | Recovery-definition sensitivity; acute node-split landmark 12 versus 26 weeks (chronic fixed at 54); scale reconciliation; clinical-unit sanity checks; transitivity/exchangeability tables; and direct-versus-indirect node splitting. |

**Table 3.**
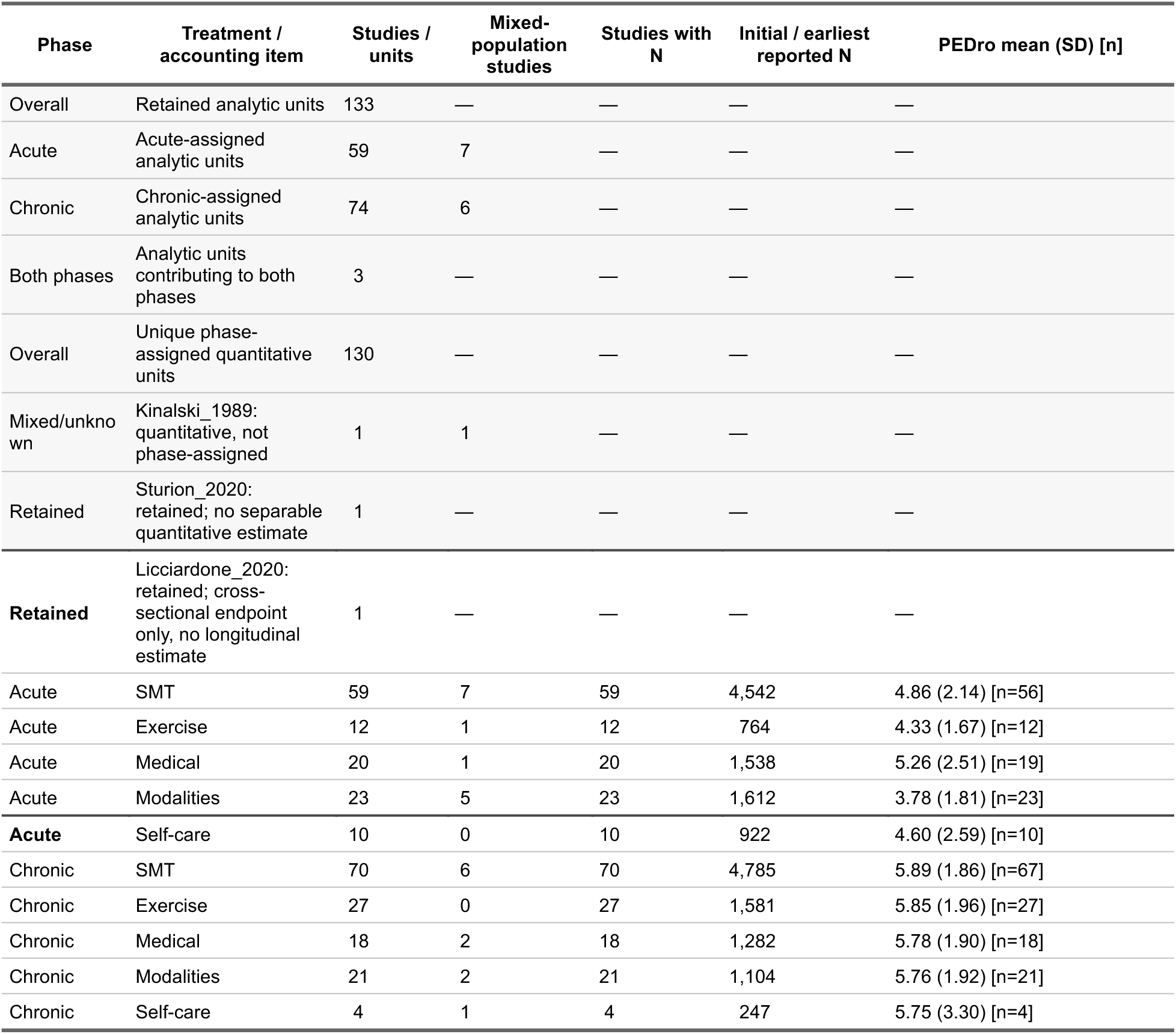
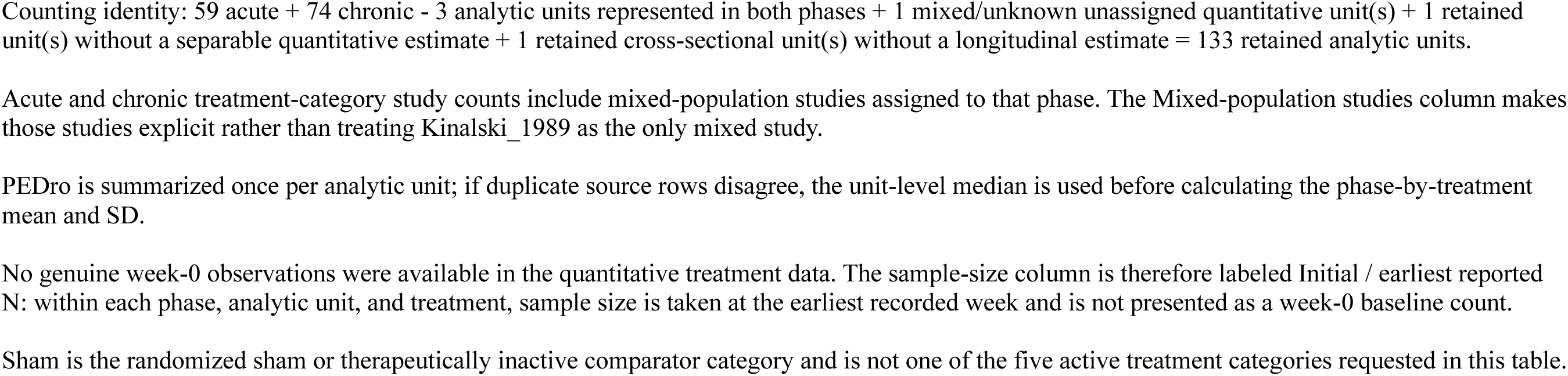
Study counts and characteristics of the comparative evidence base. The retained evidence inventory is reconciled first; treatment categories are then summarized separately by pain phase.

### Literature search for treatment studies

We searched PubMed/MEDLINE, Google Scholar, and the Cochrane Database of Systematic Reviews. We also used resources from the University of Arizona Health Sciences Library.

Searches combined terms for LBP with terms for spinal manipulation and related manual treatments. To find trials, we added terms for randomized or controlled clinical trials. We also checked the reference lists of included studies and relevant systematic reviews.

The main PubMed MeSH terms were *Manipulation, Spinal*, *Manipulation, Chiropractic*, *Manipulation, Osteopathic*, *Musculoskeletal Manipulations*, *Low Back Pain*, *Back Pain*, *Lumbosacral Region*, and *Sciatica*. We added free-text terms: spinal manipulation, chiropractic manipulation, osteopathic manipulation or osteopathic manipulative treatment (OMT), manual therapy, mobilization/mobilisation, spinal adjustment, thrust, high-velocity low-amplitude or HVLA manipulation, flexion-distraction, low back pain, lumbar pain, lumbago, lumbosacral pain, and sciatica. Trial terms were randomized/randomised, controlled clinical trial, placebo, sham, trial, and groups. In the Cochrane Central Register of Controlled Trials (CENTRAL), we did not add a trial filter, because that database contains only controlled trials. We adapted the search terms to each database’s syntax and indexing.

The evidence base was built up over time. We added studies through citation searching and relevant systematic reviews. The exact wording of every early search was not saved. The search strategy described here therefore records the concepts we used to build and update the database. It does not claim that one identical search string was run at every stage. The treatment evidence included studies of acute LBP[39, 42–99] and chronic LBP[15, 40, 41, 45, 49, 100–163]. Studies that contributed to both phases are listed in both groups.

#### Search criteria in brief

Databases: PubMed/MEDLINE, the Cochrane Central Register of Controlled Trials (CENTRAL), the Cochrane Database of Systematic Reviews, and Google Scholar, supplemented by University of Arizona Health Sciences Library resources and backward and forward citation searching of included studies and systematic reviews. Search structure: (low back pain terms) AND (spinal manipulation and manual-therapy terms) AND (randomized or controlled trial terms), using the MeSH and free-text terms listed above; no trial filter was applied in CENTRAL. Period: searches were run repeatedly between 2009 and 2026 as the evidence base was built and updated. Eligible studies: randomized or controlled clinical trials of adults with acute or chronic low back pain that included spinal manipulation or another conservative treatment (exercise, medical care, physical modalities, self-care) or a sham or inactive comparator, and that reported pain intensity at baseline and at least one follow-up in a form convertible to the common standardized scale, with arm sample sizes. Analysis windows: acute observations up to 26 weeks and chronic observations up to 54 weeks. Prospective observational cohorts were eligible only for information on change over time, as described below.

### Eligibility and study classification

We included randomized or controlled clinical trials of adults with LBP. When they added useful information, we also included prospective observational cohorts. Every study had to report a pain outcome we could convert to our common scale. Treatment arms were sorted into six categories: spinal manipulation therapy (SMT), exercise, medical care, physical modalities, self-care, and sham. Sham meant a placebo procedure or a treatment known to be ineffective. We classified each arm by what was done, not by who did it. So manipulation by a chiropractor, osteopath, physician, or physiotherapist counted as SMT if it included spinal manipulation or a similar thrust procedure. Multi-part osteopathic manipulative treatments stayed in the SMT category.

Where needed, we flagged them separately for sensitivity and transitivity checks.

Observational studies were used only for purposes their design could support. We assumed their data on how pain changes over time could stand in for the trial populations, once we accounted for clinical phase, follow-up time, differences between studies, and measured design features.

Their outcomes also had to fit our common recovery scale, and the timing of their measurements had to be clear. These assumptions are about how pain changes over time. They are not about who received which treatment. So we did not use observational cohorts to estimate treatment effects when there was no proper comparison group. Nor did we treat a treated cohort as if it were untreated natural history. Ailliet (2018) and Leemann (2014) are two prospective cohorts kept for their data on change over time, under these limits [15, 49]. Ailliet (2018) enrolled patients with neck pain or LBP; we extracted only the LBP subgroup (n = 295), using class-size– weighted means across its four LBP trajectory classes at baseline, 26 weeks and 52 weeks.

Some studies enrolled both acute and chronic patients. We kept these when the two groups could be separated. Acute data went into the acute analysis and chronic data into the chronic analysis. If a study’s phase could not be assigned reliably, it stayed in the inventory but was left out of both models. Studies were excluded only because of their population, treatment, design, missing outcomes, or data that could not be mapped to our outcome. They were never excluded because of the direction or size of their results.

### Outcome extraction and standardization

Pain was the main outcome. Numerical rating scale (NRS) and visual analogue scale (VAS) scores were converted to a common 0–100 scale before we calculated standardized change. Many studies measured pain more than once during follow-up. We kept each follow-up as a separate time point within its original arm or cohort. We did not count repeated measurements as extra arms or extra patients [164].

To describe how severe pain was at enrollment, we also put baseline pain on a 0–10 scale. Scores already on a 0–10 scale were kept. Scores on a 0–100 scale were divided by 10. Each physical arm’s baseline was counted only once. Arm means were weighted by the number of patients enrolled to get summaries by study, treatment, and phase. This baseline description was not used to define treatment effects.

Improvement was expressed as Hedges’ *g* [165]. This is the change in pain divided by the pooled baseline standard deviation. It puts studies that used different pain scales on the same footing.

Positive values mean improvement. Each study kept its identifier throughout. That way, repeated measurements and multiple arms from one study were not treated as if they came from separate studies.

We also converted improvement into the chance of recovery, because that is easier to interpret clinically. Our main definition of recovery was pain of 2/10 or less. The conversion was calibrated separately for each phase. Average starting pain, weighted by sample size, was 4.823/10 in acute LBP and 5.284/10 in chronic LBP. We used a fixed standard deviation of 2.0 NRS points (SD_NRS = 2.0). To reach 2/10, patients therefore needed to drop 2.823 points (acute) or 3.284 points (chronic). In standardized units, these thresholds are Δ = 1.412 (acute) and Δ = 1.642 (chronic). The chance of recovery was P(recovery) = Φ(*g* − Δ_phase), where Φ is the standard normal cumulative distribution function [166, 167]. This is a model-based translation of average improvement into a recovery rate. It is not a directly counted proportion of recovered patients. The baseline means and the SD were treated as fixed values, not as uncertain quantities. The 2/10 threshold was set in advance [168]. Chronic LBP was handled differently.

Paper I’s chronic natural-history model estimates the probability of a favorable course at 54 weeks, using its reference (DIRECT) outcome family. A favorable course is not the same as pain of 2/10 or less. So for chronic LBP the natural-history posterior entered the treatment model directly, as its starting line on the *g* scale,

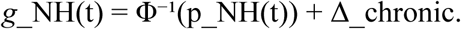

Each treatment’s recovery is then

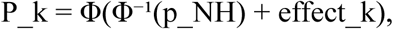

where effect_k is that treatment’s standardized deviation from natural history. In chronic LBP, “recovery” therefore means a favorable course. We also tested three other definitions of recovery: any improvement, at least 30% reduction, and at least 50% reduction.

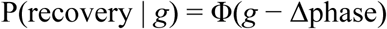

For each posterior draw s, recovery and the treatment-minus-sham difference are calculated as follows:

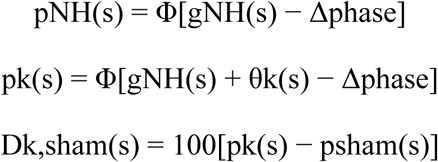

where Φ is the standard normal cumulative distribution function; *g* is standardized mean improvement; Δphase is the phase-specific standardized recovery threshold (acute 1.412; chronic 1.642); s indexes posterior draws; gNH(s) is the natural history value in draw s; pNH(s), pk(s), and psham(s) are the recovery probabilities for natural history, treatment k, and sham, respectively; θk(s) is the standardized treatment effect for treatment k in draw s; and Dk,sham(s) is the treatment-minus-sham difference in recoveries per 100 persons.

In plain terms, *g* is how far the average patient in an arm has moved, in baseline standard deviations. Δ is how far a patient must move to cross the recovery line, from the average starting pain down to 2/10. Suppose individual changes are roughly bell-shaped around the arm average, with the same SD used for standardizing. Then Φ(*g* − Δ) is the share of patients expected to cross the line. In acute LBP, Δ = 1.412. An arm that improves by *g* = 1.0 (about 2 NRS points) gives Φ(−0.41) ≈ 34% recovered. An arm that improves by *g* = 0.5 gives about 18%. The threshold is essential. Without it, Φ(*g*) would count almost any average improvement as recovery. For example, *g* = 1.0 would give 84%, turning a modest effect into near-certain success. Because the conversion relies on a distributional assumption, it is a model-based estimate, not an observed count. The four recovery definitions in the sensitivity analysis show how much the conclusions depend on it [166, 167].

The natural-history and treatment posteriors come from separate evidence. So we paired their draws at random to simulate their joint distribution (Monte Carlo sampling). We calculated posterior means and credible intervals only after converting each draw to a recovery probability.

The order matters because Φ is a curve, not a straight line. The average of converted values is not the same as converting the average. Suppose natural-history uncertainty came down to just two equally likely draws: *g* = 0 and *g* = 1.0. In acute LBP, these give recovery rates of 7.9% and 34.0%, which average 21%. If we average *g* first (0.5) and then convert, we get 18%. Converting each draw and then summarizing keeps the full uncertainty in the result. Converting a single point estimate quietly throws it away [169]. Pairing the draws at random is valid here because the two posteriors share no data. It would be wrong if the same trials had informed both.

## Study quality

Each randomized study received a single rating on the PEDro quality scale. Ratings were made with the assistance of an artificial intelligence model (ChatGPT). They were not duplicated and were not checked against ratings by human reviewers.

PEDro was designed for randomized trials. For nonrandomized studies, we therefore described design and source separately rather than forcing them onto the PEDro scale. PEDro entered both treatment models as a standardized study-level covariate, so its coefficient is the change in improvement per 1 SD of PEDro score; studies without a score were set to the mean. PEDro was also used in sensitivity analyses. It was not used to exclude studies.

## External natural-history posterior from linked Paper I

Paper I estimated natural history on its own, using separate prognosis studies. Paper I defines the outcomes for each phase, the sources, how chronic outcomes were judged, the Bayesian models, the validation checks, and the uncertainty. This paper imports those results unchanged. The natural-history posteriors from Paper I enter this paper as external prior distributions for the background trajectory. They are not updated by the treatment trials; their uncertainty is propagated into every treatment estimate. An audit confirmed that no treatment trial fed into the Paper I model. It also confirmed that no prognosis cohort was treated as an arm in our network. The 26-week (acute) and 54-week (chronic) time points are when we compare treatments. They do not limit which prognosis studies Paper I could use.

## Treatment effects relative to natural history and sham

For each phase, we modeled the standardized improvement in each treatment arm. The model accounts for four sources of variation. These are the uncertainty within each observation, extra unexplained scatter, differences between studies, and the structure of the treatment network. At the final step, we paired draws from the Paper I natural-history posterior with draws from the treatment posterior. Natural history is therefore an outside reference for “what would have happened anyway.” It is not an extra node in the network. We formed a recovery comparison only after confirming that natural history and treatment measured the same outcome, or were linked by a measurement model set out in advance.

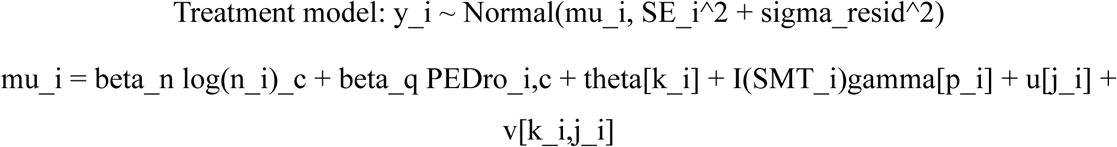

where y_i is standardized within-arm improvement for observation i; mu_i is its treatment-model expectation; SE_i is its within-observation standard error; theta[k] is the treatment-category effect; gamma[p] is an SMT-provider deviation where applicable; u[j] is a study random effect; and v[k,j] is a treatment-by-study deviation. The independent natural-history posterior is introduced only after this treatment model has produced posterior draws.

Each term in the model soaks up a different source of variation, so that it is not mistaken for a treatment effect. θ[k] is the quantity we care about: the average improvement for treatment category k. u[j] lets each study sit higher or lower overall. This reflects its patients, setting, and measures. All arms in a study share u[j], so comparisons within a study are not affected by it. v[k,j] lets a treatment do somewhat better or worse in one study than on average. This is how the model captures trial-to-trial variation in the treatment effect itself. γ[p] lets SMT delivered by different types of provider differ from the SMT average. The log(n) and PEDro terms adjust for patterns linked to small studies and to study quality. SE_i² carries each observation’s own sampling uncertainty. σ_resid² adds variance when the data scatter more than their reported standard errors predict. As in the chronic natural-history model of Paper I, these random effects let studies differ without letting those differences pose as treatment effects.

We used two reference comparisons. Treatment versus natural history compares modeled recovery on treatment with the independently estimated course of untreated LBP. Treatment versus sham compares one treatment with sham inside the network. These answer different questions. They are not parts that add up to a total. We estimated placebo responsiveness as sham minus natural history, which removes the recovery expected anyway.

## Placebo responsiveness = sham − natural history

We ran the network meta-analysis separately for the acute and chronic networks, using the six treatment categories. Trials with more than two arms kept their within-study structure. We report posterior means and 95% credible intervals (CrIs). A standardized effect of about ±0.30 served as a rough guide to clinical importance. It was a reference zone, not a pass/fail test of significance [168, 170].

## Absolute-recovery benchmark and posterior decision probability

For each treatment, we asked: in what share of posterior draws did the modeled recovery rate exceed 50%? This tells us how sure we are about the population recovery rate, given the model, the outcome definition, and the follow-up time. It is not the chance that a given patient benefits from treatment. It is not the share of patients whose improvement was caused by treatment. And it is not the size of the treatment effect. The 50% benchmark is also different from three other cut-offs used in this paper: the individual recovery definition (pain ≤2/10), the 50% pain-reduction definition used in sensitivity analyses, and the ±0.30 reference zone for effect size. The benchmark is not a validated policy threshold. We calculated these probabilities from the saved posterior draws at each phase’s endpoint.

Two probabilities are easy to confuse here. The chance that the population recovery rate exceeds 50% describes our certainty. It does not describe patients. Imagine a treatment with a modeled recovery of 55% and a narrow interval. Its chance of exceeding the benchmark could be above 90%. Yet only about 55 of 100 patients would be expected to recover. The posterior says nothing about which patients recover, or whether treatment caused it. In contrast, a treatment with a modeled recovery of 45% and a wide interval might have a 30% chance of exceeding the benchmark (hypothetical values). The first number is confidence about a population value. The second is the population value itself.

## Transitivity, exchangeability, and network assumptions

We checked transitivity and exchangeability separately in the acute and chronic networks. (Both are explained below.) We compared possible effect modifiers across treatment comparisons.

These included clinical phase, follow-up time, baseline pain, sample size, study quality, treatment category, provider type where known, and other available study features. We displayed these checks as grids of plots (lattices) for each phase. This let us look for imbalance across many features at once, rather than judging one covariate at a time.

We also assessed exchangeability through differences between studies, study-level random effects, residual patterns, the link between treatment and quality, and sensitivity analyses. Where studies overlapped little on these features, we interpreted direct and indirect evidence with caution. These checks show whether the assumptions are plausible. They do not prove that transitivity holds.

An indirect comparison works through a shared comparator. Suppose SMT beats sham by *g* = 0.2 in one set of trials, and exercise beats sham by *g* = 0.1 in another. The network then infers that SMT beats exercise by about 0.1 (hypothetical values). That inference holds only if both sets of sham-controlled trials enrolled similar patients. Only then is sham the same yardstick in each. If the SMT trials had enrolled milder or more recent cases, the indirect estimate would compare patient groups as well as treatments. Transitivity is the assumption that the yardstick is shared.

Exchangeability is the closely related assumption that the trials are similar enough to be combined. The lattice plots check whether measured effect modifiers are spread evenly enough across comparisons for these assumptions to be plausible [171].

## Direct and indirect evidence as an NMA validity check

Before interpreting the network estimates, we checked whether the evidence could support valid indirect comparisons. We first confirmed that the network was connected and reviewed the transitivity and exchangeability checks described above. We then used standard random-effects NMA diagnostics. These looked at differences between studies, overall inconsistency, and local agreement between direct and indirect evidence. We checked local inconsistency by node splitting. This compares the effect estimated from head-to-head trials with the effect estimated indirectly through the rest of the network [171–173].

We treated these checks as a requirement for interpreting the NMA, not as optional extras. We looked closely at how large and how uncertain any direct–indirect disagreement was. We also noted whether each comparison rested mainly on direct or indirect evidence. A non-significant inconsistency test was not taken as proof of consistency, especially when a comparison had little data. Where direct and indirect estimates disagreed, or where effect modifiers overlapped little, we interpreted the network estimate more cautiously.

Take modalities versus SMT as an example. The network has two kinds of evidence for this pair.

Some trials compared the two head-to-head. Other evidence comes indirectly, through other treatments. Node splitting estimates the comparison from each source separately and asks whether they agree. Agreement supports combining them. Disagreement suggests that some trials differ in ways that break the shared-yardstick assumption. When a comparison has little data, both routes give wide intervals. So failing to find disagreement is only weak evidence that the two sources agree [174].

## Leakage and independence audit

We brought in natural history on purpose as outside evidence. So we audited study names and sources to make sure no study appeared in both the natural-history set and the treatment network. We checked exact names and sources to catch duplicates. Reviews that included cohorts already used in the natural-history model were not added as extra natural-history studies. Sham arms from the treatment network were not used to build the natural-history trajectory. This kept information from the treatment trials out of the benchmark used to judge those same trials.

Independence matters for a simple reason. If the same trials had helped build the natural-history benchmark and were then judged against it, the benchmark would be partly made from the results it is meant to test. Those trials would be pulled toward agreeing with it. Treatment-versus-natural-history differences would then shrink toward zero by design. This is the same error as using one dataset both to pick a hypothesis and to test it, sometimes called “double dipping” [175]. Keeping the two evidence streams apart means the benchmark was fixed before we looked at the treatment data.

## Sensitivity and validation analyses

Sensitivity analyses tested other natural-history models, the recovery threshold, assumptions about treatment rates, study quality, blinding, and the line between natural history and sham-related improvement. Common-asymptote checks asked whether, in acute LBP, treatments might differ only in how fast patients reach the same end point. This check assumed a shared Bayesian hierarchical natural-history end point, with study as a random factor.

The main sensitivity analyses covered: the definition of recovery; two methods for translating treatment time (model-based versus the older method); node-splitting at 12 versus 26 weeks in acute LBP (chronic stayed at 54 weeks); reconciling scales; sanity checks in clinical units; and meta-regression on study quality and blinding. We treated the PEDro-based blinding analysis as a rough proxy for overall study quality. That is because PEDro combines blinding with allocation, dropout, intention-to-treat, and other design features.

Paper I reports a linear-form sensitivity analysis for the acute natural-history trajectory; under that form, natural recovery at 26 weeks was 79.9%, compared with 75.2% under the main saturating (leveling-off) form. We therefore reran the full acute treatment model using the linear-form natural-history posterior. This rerun used the same 220 treatment observations, 46 studies, and treatment categories as the main model, so it isolates the effect of curve shape. We compared treatment-versus-sham conclusions directly with the main model (Table 9).

## Bayesian computation and convergence

Models were fitted by Markov chain Monte Carlo (MCMC) in JAGS, with four chains per model [176]. All preset convergence criteria were met before results were carried forward. The maximum potential scale reduction factor (PSRF) was 1.0007 (acute) and 1.0026 (chronic) for the treatment models, and 1.0034 and 1.0029 for the natural-history models [177, 178]. Values this close to 1.0 mean the chains agreed. Iteration counts, thinning, minimum effective sample sizes, and retry rules are in Table 2.

## Results

### Evidence base

The final evidence base had 133 analytic units from 131 parent trials. Fifty-nine units went into the acute analysis and 74 into the chronic analysis. Three units gave separable data to both. That leaves 130 unique units assigned to a phase. Studies that enrolled both acute and chronic patients were kept when the two groups could be separated. Their data went into the right analysis instead of being dropped as mixed. Kinalski (1989)[39] was the only unit whose phase could not be assigned, so it entered neither model. Sturion (2020)[40] stayed in the inventory but gave no separate treatment estimate. Licciardone (2020) reported only a single cross-sectional pain score, so it also stayed in the inventory without a longitudinal estimate. These categories add up exactly to the 133 units. Table 1 summarizes treatments, Table 2 specifies the model, and Table 3 accounts for every unit by phase.

Both treatment networks were connected. That is, every treatment could be linked to every other through some chain of trials. In the network display, SMT had 68 acute and 74 chronic arms.

Head-to-head evidence was strongest for modalities versus SMT (23 analytic study units) and medical care versus SMT (20) in acute LBP. In chronic LBP, it was strongest for sham versus SMT (27), exercise versus SMT (24), modalities versus SMT (20), and medical care versus SMT (18). Figure 1 shows the network, and Table 4 summarizes the head-to-head evidence.

**Figure 1.**
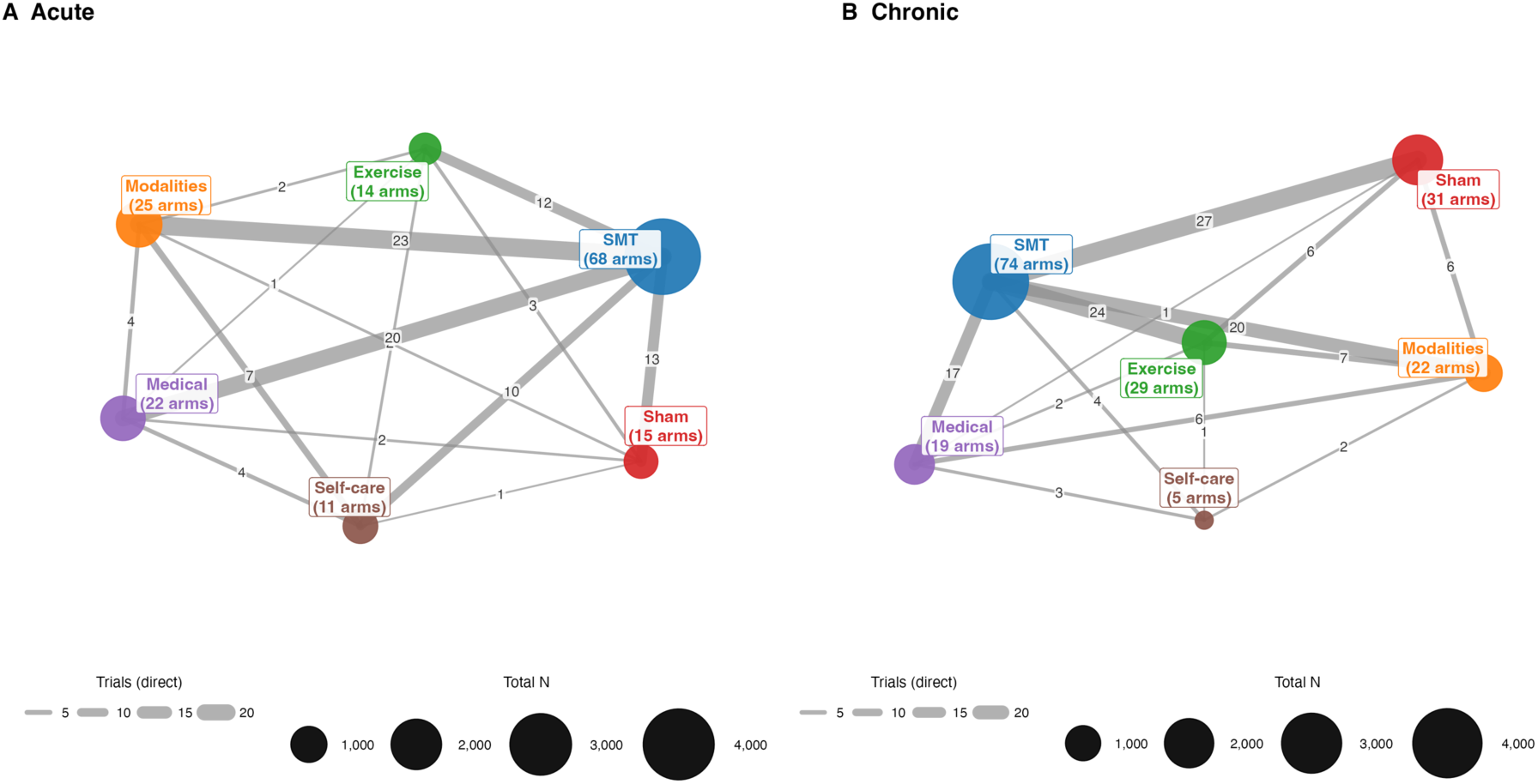
Treatment evidence networks for acute and chronic low back pain. Node size reflects the number of patients at the first reported time point. Node labels give the number of arms. Line width and line labels give the number of analytic units that compared each pair of treatments head-to-head. Sham is the comparator category. Arms from multi-arm studies can contribute to more than one pair.

**Table 4.** Direct comparative evidence supporting the network.

| Comparison | Contributing analytic study units |
| --- | --- |
| **ACUTE** |  |
| Modalities vs SMT | 23 |
| Medical vs SMT | 20 |
| Sham vs SMT | 13 |
| Exercise vs SMT | 12 |
| Self-care vs SMT | 10 |
| Modalities vs Self-care | 7 |
| Medical vs Modalities | 4 |
| Medical vs Self-care | 4 |
| Sham vs Exercise | 3 |
| Exercise vs Modalities | 2 |
| Exercise vs Self-care | 2 |
| Sham vs Medical | 2 |
| Sham vs Modalities | 2 |
| Exercise vs Medical | 1 |
| Sham vs Self-care | 1 |
| **CHRONIC** |  |
| Sham vs SMT | 27 |
| Exercise vs SMT | 24 |
| Modalities vs SMT | 20 |
| Medical vs SMT | 17 |
| Exercise vs Modalities | 7 |
| Medical vs Modalities | 6 |
| Sham vs Exercise | 6 |
| Sham vs Modalities | 6 |
| Self-care vs SMT | 4 |
| Medical vs Self-care | 3 |
| Exercise vs Medical | 2 |
| Modalities vs Self-care | 2 |
| Exercise vs Self-care | 1 |
| Sham vs Medical | 1 |

Multi-arm analytic study units can contribute to more than one treatment pair but are counted once per pair.

### NMA validity: direct and indirect evidence

Before interpreting treatment effects, we checked whether each network met the assumptions needed to combine direct and indirect evidence. Both networks were connected. Measured effect modifiers were similar enough across comparisons to support exchangeability, but only conditionally, not without limits. We then used standard random-effects diagnostics to look at differences between studies, overall inconsistency, and local agreement between direct and indirect evidence [171–173, 179].

We examined node-splitting results one comparison at a time, rather than relying on P values alone. Comparisons with many trials rested mostly on head-to-head evidence. Comparisons with few trials leaned more on indirect routes through shared comparators. We used the size and uncertainty of any disagreement to temper our interpretation. Figure 2 shows each comparison that had both direct and indirect evidence. Table 4 gives the number of analytic units behind each head-to-head pair. The standard 12-week acute network showed moderate heterogeneity (τ ≈ 0.13; I² ≈ 27%). The chronic 54-week network showed more (τ ≈ 0.34; I² ≈ 73%). Here τ is the standard deviation of true effects across studies, and I² is the share of total variation due to real differences between studies rather than chance. Both point to caution when reading individual comparisons.

**Figure 2.**
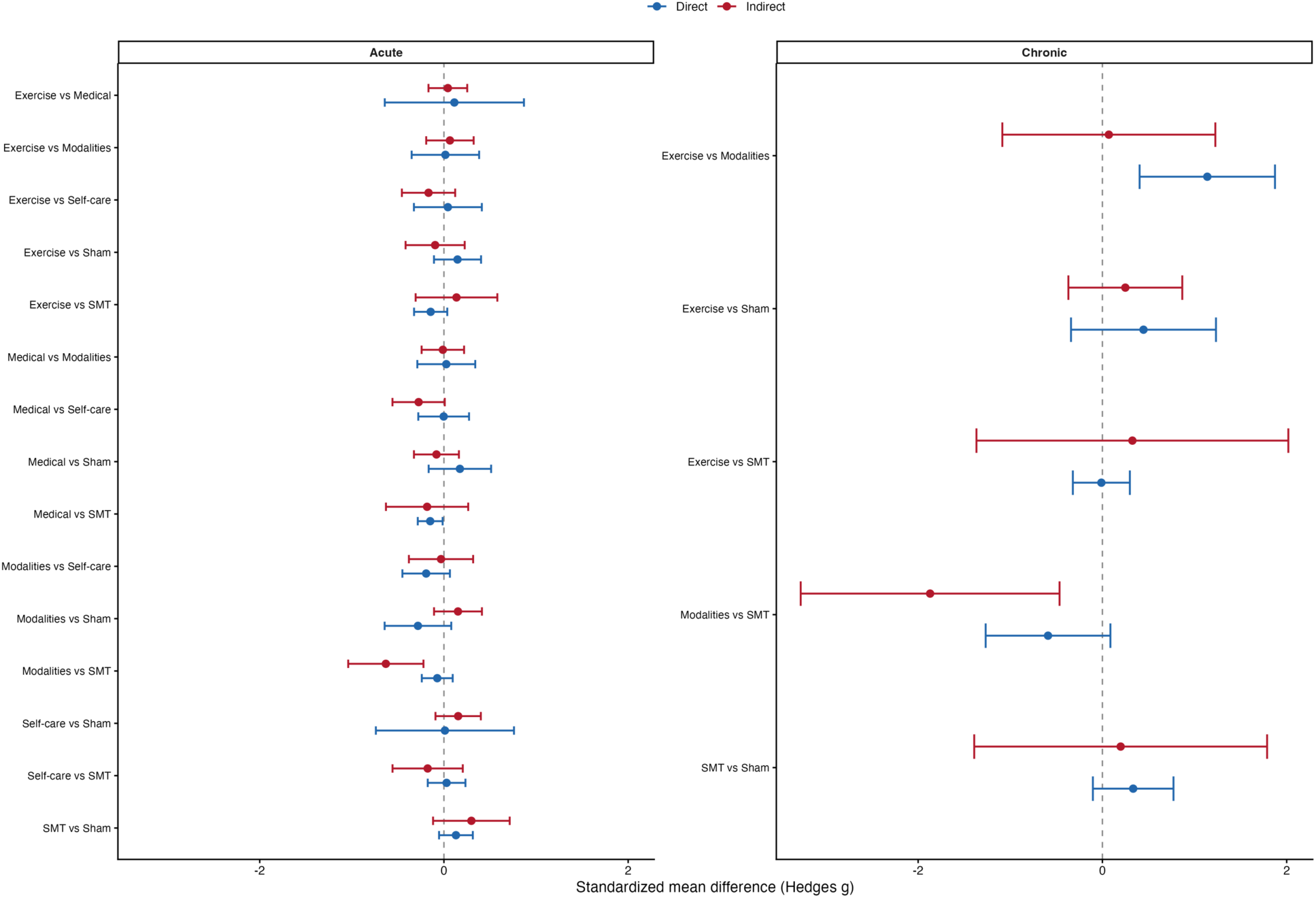
Direct versus indirect evidence for treatment comparisons. Random-effects node splitting compares direct and indirect estimates at preset time points for each phase. Only comparisons with both kinds of evidence are shown. We consider the size and uncertainty of any disagreement, not P values alone.

### Natural-history trajectories

The natural-history estimates come from Paper I. The acute estimate drew on 8 studies (26 observations). The chronic estimate drew on 12 cohorts (15 observations). Table 5 lists the sources. Modeled natural-history recovery was 75.2% (95% CrI 49.8% to 94.3%) at 26 weeks in acute LBP. In chronic LBP, the probability of a favorable course at 54 weeks was 18.1% (8.8% to 31.9%).

**Table 5.** Natural History source studies and provenance.

| Cohort | Source / provenance | Role | Reason / use |
| --- | --- | --- | --- |
| **ACUTE** |  |  |  |
| Chapman 2012 | external natural-course/prognosis cohort | primary | Standardized pain change on the common scale |
| Chen 2007 | external natural-course/prognosis cohort | primary | Standardized pain change on the common scale |
| Epping-Jordan 1998 | external natural-course/prognosis cohort | primary | Standardized pain change on the common scale |
| Coste 1994 | inception cohort | primary | Direct recovery observations |
| Grotle 2005 | primary-care prognosis cohort | primary | Direct recovery observations |
| Henschke 2008 | inception cohort | primary | Direct complete-recovery observations |
| Schiøttz-Christensen 1999 | general-practice prognosis cohort | primary | Functional recovery is primary endpoint; complete recovery retained for sensitivity |
| Oliveira 2021 | emergency-department inception cohort | primary | Usual-care prognosis cohort; directly reported complete recovery |
| Ferguson 2000 | prospective natural-course cohort | validation | Excellent trajectory validation but reported functional-performance probabilities are not the same estimand as patient recovery probability |
| van den Hoogen 1998 | mixed-duration general-practice cohort | validation | 77% recent onset and 23% chronic; subgroup-specific recovery probabilities not extractable, so not used to define either phase curve |
| Wahlgren 1997 | subacute transition cohort | validation | Selected at 6-10 weeks; enriched for persistence; transition validation only |
| Pengel 2003 | systematic review | source_map | Source map and external validation; not entered as an independent study |
| Costa 2012 pooled | meta-analysis | validation | Meta-analytic validation only; component cohorts are preferred for fitting |
| **CHRONIC** |  |  |  |
| Costa 2009 | external chronic natural-course cohort | primary | STRICT family: Pain-free for 30 consecutive days (39.1, 52.2 wk) |
| Tamcan 2010 | external chronic natural-course cohort | primary | COURSE family: Fluctuating chronic/recurrent trajectory (52 wk) |
| Dunn 2006 | external chronic natural-course cohort | primary | DIRECT family: Recovering trajectory class (26 wk) |
| Von Korff 1993 | external chronic natural-course cohort | primary | STRICT family: Pain-free at one year (52 wk) |
| Carey 2000 | external chronic natural-course cohort | primary | COURSE family: Remitting rather than unremitting course (95.7 wk) |
| van den Hoogen 1998 | external chronic natural-course cohort | primary | COURSE family: Ended episode without reported relapse (52 wk) |
| Biering-Sørensen 1983 | external chronic natural-course cohort | primary | DIRECT family: Less pronounced course - men; Less pronounced course - women (52 wk) |
| Waxman 2000 | external chronic natural-course cohort | primary | COURSE family: Resolved annual LBP pattern (156.6 wk) |
| van Oostrom 2011 | external chronic natural-course cohort | primary | DIRECT family: Recovery pattern across repeated waves (521.8 wk) |
| Nordstoga 2017 (HUNT) | external chronic natural-course cohort | primary | STRICT family: No chronic-LBP criterion - women; No chronic-LBP criterion - men (573.9 wk) |
| Gomes 2023 (EpiDoC) | external chronic natural-course cohort | primary | COURSE family: Relapsing rather than persistent course (260.9 wk) |
| Rabiei 2026 | external chronic natural-course cohort | primary | DIRECT family: Gradually improving pain trajectory (104.4 wk) |
| Wallwork 2024 | systematic review/meta-analysis | validation | Validation curve only; excluded from fit to avoid double counting component primary cohorts |
| McGorry 2000 | 6-month nonintervention diary cohort | validation | Strong evidence for episodic chronic/recurrent course but no common recovery-scale longitudinal aggregate suitable for primary fit |
| Von Korff 1996 | review | source_map | Review/source map; not an independent cohort |

Paper I models and documents how the natural-history studies differ from one another. This paper takes the resulting posterior as given and does not refit the prognosis data. Figure 3 shows the Paper I natural-history curves alongside the treatment curves.

**Figure 3.**
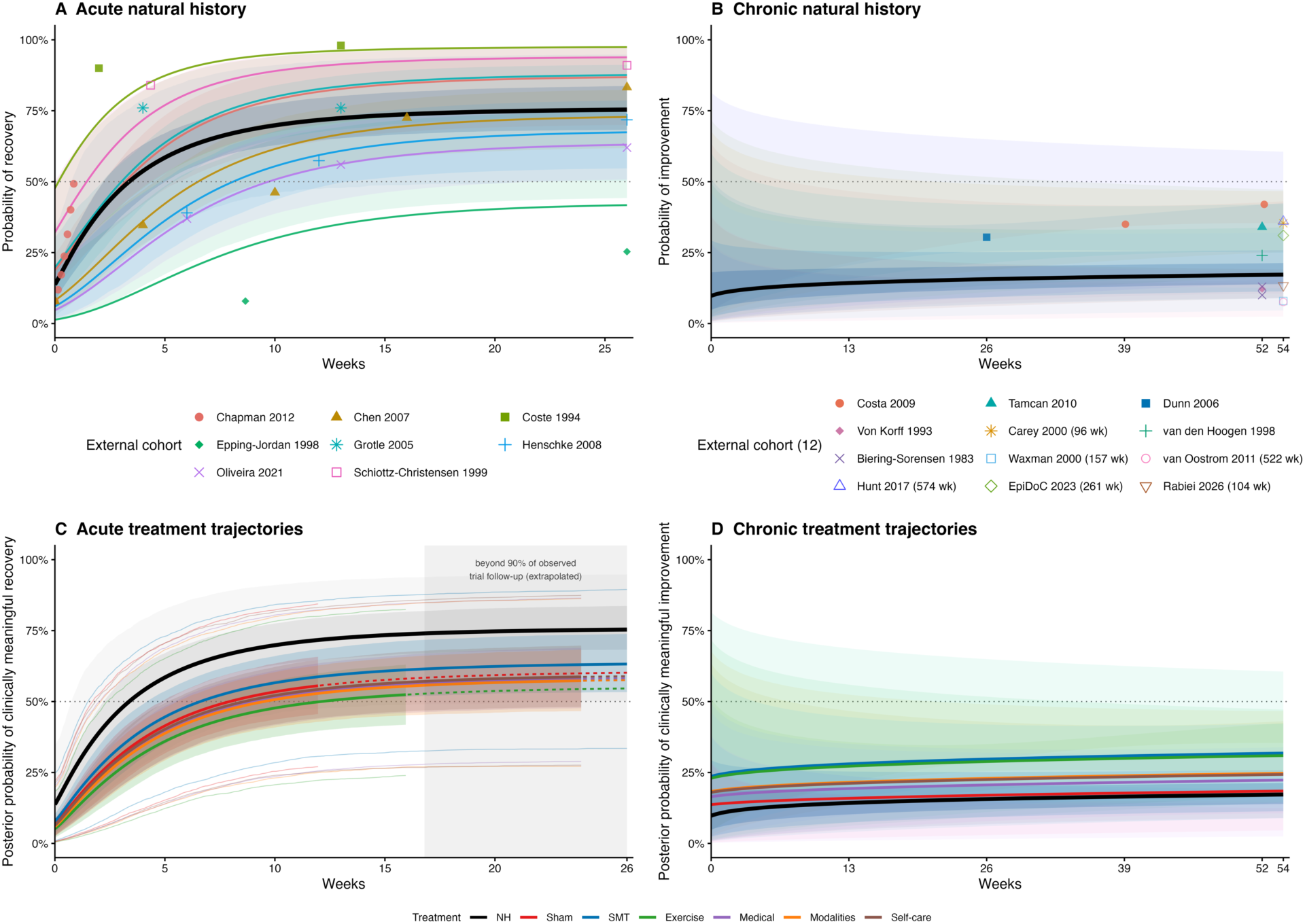
Natural-history and treatment trajectories. Natural-history curves come from Paper I. Treatment curves come from the network models for each phase. In panel C, each treatment’s curve is solid over the weeks covered by at least two of its trials (sham to 12 weeks, exercise to 16, medical care, modalities and self-care to 24, and SMT to 26) and dotted beyond; the shaded region lies beyond 90% of acute trial follow-up. In panel D, all chronic treatments are drawn to 54 weeks; sham, SMT, exercise and modalities trials extend to 48–52 weeks, and medical care and self-care trials to about 24 weeks.

In acute LBP, no treatment arm did better than natural history. Per 100 patients, differences from natural history ranged from −12.0 for SMT (95% interval −23.2 to −0.7) to −20.7 for exercise (−33.7 to −5.3), and every interval lay below zero. Compared with sham in the network, SMT gave 3.1 more recoveries per 100 (95% interval −6.3 to +12.8). Medical care, exercise, modalities, and self-care ranged from −5.7 to −1.3 per 100. So recovery during acute treatment was high, but natural recovery was even higher. We found no clear treatment-specific signal on top of it. Table 6 gives the exact estimates.

**Table 6.** Clinical interpretation of treatment outcomes.

| Treatment | Week 12:<br>per 100 | Week 12: 95%<br>interval | Horizon:<br>per 100 | Horizon: 95%<br>interval |
| --- | --- | --- | --- | --- |
| **ACUTE — MODELED RECOVERY** |  |  |  |  |
| Sham | 55.7 | [27.4, 84.3] | 60.2 | [30.0, 88.3] |
| SMT | 58.9 | [30.7, 86.0] | 63.2 | [33.5, 89.6] |
| Medical | 53.5 | [25.7, 82.7] | 58.1 | [28.3, 86.9] |
| Exercise | 49.9 | [22.4, 80.6] | 54.5 | [24.8, 85.2] |
| Modalities | 52.9 | [25.2, 82.4] | 57.5 | [27.8, 86.6] |
| Self-care | 54.3 | [25.5, 83.8] | 58.8 | [28.1, 87.8] |
| **ACUTE — DIFFERENCE VS NATURAL HISTORY** |  |  |  |  |
| Sham | -15.9 | [-28.3, -2.5] | -15.0 | [-27.7, -2.1] |
| SMT | -12.7 | [-23.8, -0.9] | -12.0 | [-23.2, -0.7] |
| Medical | -18.0 | [-29.4, -4.3] | -17.1 | [-28.9, -3.6] |
| Exercise | -21.6 | [-34.1, -6.3] | -20.7 | [-33.7, -5.3] |
| Modalities | -18.6 | [-30.0, -4.9] | -17.7 | [-29.5, -4.1] |
| Self-care | -17.2 | [-30.8, -2.5] | -16.4 | [-30.3, -2.1] |
| **ACUTE — DIFFERENCE VS NMA SHAM** |  |  |  |  |
| Sham | 0.0 | [+0.0, +0.0] | 0.0 | [+0.0, +0.0] |
| SMT | 3.2 | [-6.5, +13.1] | 3.1 | [-6.3, +12.8] |
| Medical | -2.2 | [-12.3, +7.9] | -2.1 | [-12.1, +7.8] |
| Exercise | -5.8 | [-16.9, +5.0] | -5.7 | [-16.7, +4.9] |
| Modalities | -2.8 | [-13.0, +7.4] | -2.7 | [-12.8, +7.3] |
| Self-care | -1.4 | [-13.6, +10.6] | -1.3 | [-13.4, +10.4] |
| **CHRONIC — MODELED RECOVERY** |  |  |  |  |
| Sham | 15.8 | [4.6, 35.1] | 18.5 | [8.4, 33.6] |
| SMT | 27.9 | [10.0, 52.6] | 31.8 | [16.3, 51.3] |
| Medical | 19.3 | [5.7, 41.3] | 22.4 | [10.1, 40.1] |
| Exercise | 27.1 | [9.9, 51.3] | 30.9 | [16.5, 49.7] |
| Modalities | 21.3 | [6.8, 43.8] | 24.6 | [11.9, 42.4] |
| Self-care | 21.1 | [6.0, 45.1] | 24.4 | [10.3, 44.5] |
| **CHRONIC — DIFFERENCE VS NATURAL HISTORY** |  |  |  |  |
| Sham | 0.4 | [-3.9, +5.3] | 0.4 | [-4.2, +5.7] |
| SMT | 12.4 | [+3.3, +22.1] | 13.8 | [+4.3, +22.6] |
| Medical | 3.9 | [-2.2, +12.5] | 4.3 | [-2.5, +13.1] |
| Exercise | 11.6 | [+4.2, +20.4] | 12.9 | [+5.7, +20.9] |
| Modalities | 5.9 | [+0.2, +13.9] | 6.6 | [+0.2, +14.5] |
| Self-care | 5.6 | [-2.6, +17.8] | 6.3 | [-3.0, +18.7] |
| **CHRONIC — DIFFERENCE VS NMA SHAM** |  |  |  |  |
| Sham | 0.0 | [+0.0, +0.0] | 0.0 | [+0.0, +0.0] |
| SMT | 12.0 | [+3.4, +21.3] | 13.4 | [+4.3, +21.9] |
| Medical | 3.5 | [-3.4, +12.2] | 3.9 | [-3.9, +12.9] |
| Exercise | 11.2 | [+4.1, +20.0] | 12.5 | [+5.5, +20.4] |
| Modalities | 5.5 | [-0.4, +13.4] | 6.2 | [-0.5, +13.9] |
| Self-care | 5.3 | [-3.5, +17.2] | 5.9 | [-4.0, +18.1] |

Every acute arm, sham included, fell below natural history. It is unlikely that every treatment, including sham, slows recovery. A more likely explanation is that trial patients and natural-history cohorts differ in systematic ways. Trials enroll people who seek care and who meet severity and duration criteria, often during a flare. Trial eligibility rules, such as a minimum pain score, can also exclude people with the mildest pain, who tend to recover fastest; inception and community cohorts include them [180]. Differences in when and how outcomes are measured can add to this gap. The treatment-versus-natural-history comparison therefore mixes any treatment effect with a difference between populations. That is one reason we also report treatment versus sham within the network, where this problem does not arise. The similar pattern of improvement across very different treatments matches earlier findings [7].

The comparisons with natural history at the reporting horizons also rest on a modeling assumption. The model treats each treatment’s effect as a constant shift from the natural-history curve. Yet the acute trial observations average only 7.1 weeks of follow-up, and the chronic observations 13.1 weeks against a 54-week horizon. Estimates at 26 and 54 weeks therefore carry that shift beyond most of the trial data; in Figure 3, panel C, each acute curve turns dotted where its trials end. Comparisons with sham are less affected, because sham and active arms share the same time structure. A sensitivity analysis that lets treatments change the rate of recovery, rather than shift it, is reported below.

Modeled recovery and differences versus Natural History and NMA sham, per 100 patients, at 12 weeks and at the reporting horizon (acute 26 weeks; chronic 54 weeks).

Using sham instead of natural history as the comparator led to the same conclusion for acute LBP. Standardized treatment-versus-sham effects were small and uncertain: SMT *g* = 0.09 (95% CrI −0.18 to 0.35), exercise *g* = −0.16 (−0.45 to 0.13), modalities *g* = −0.07 (−0.35 to 0.20), self-care *g* = −0.04 (−0.36 to 0.28), and medical care *g* = −0.06 (−0.33 to 0.21). Figure 4 shows these standardized effects alongside the same results per 100 patients. Table 7 gives the full acute model summary.

**Figure 4.**
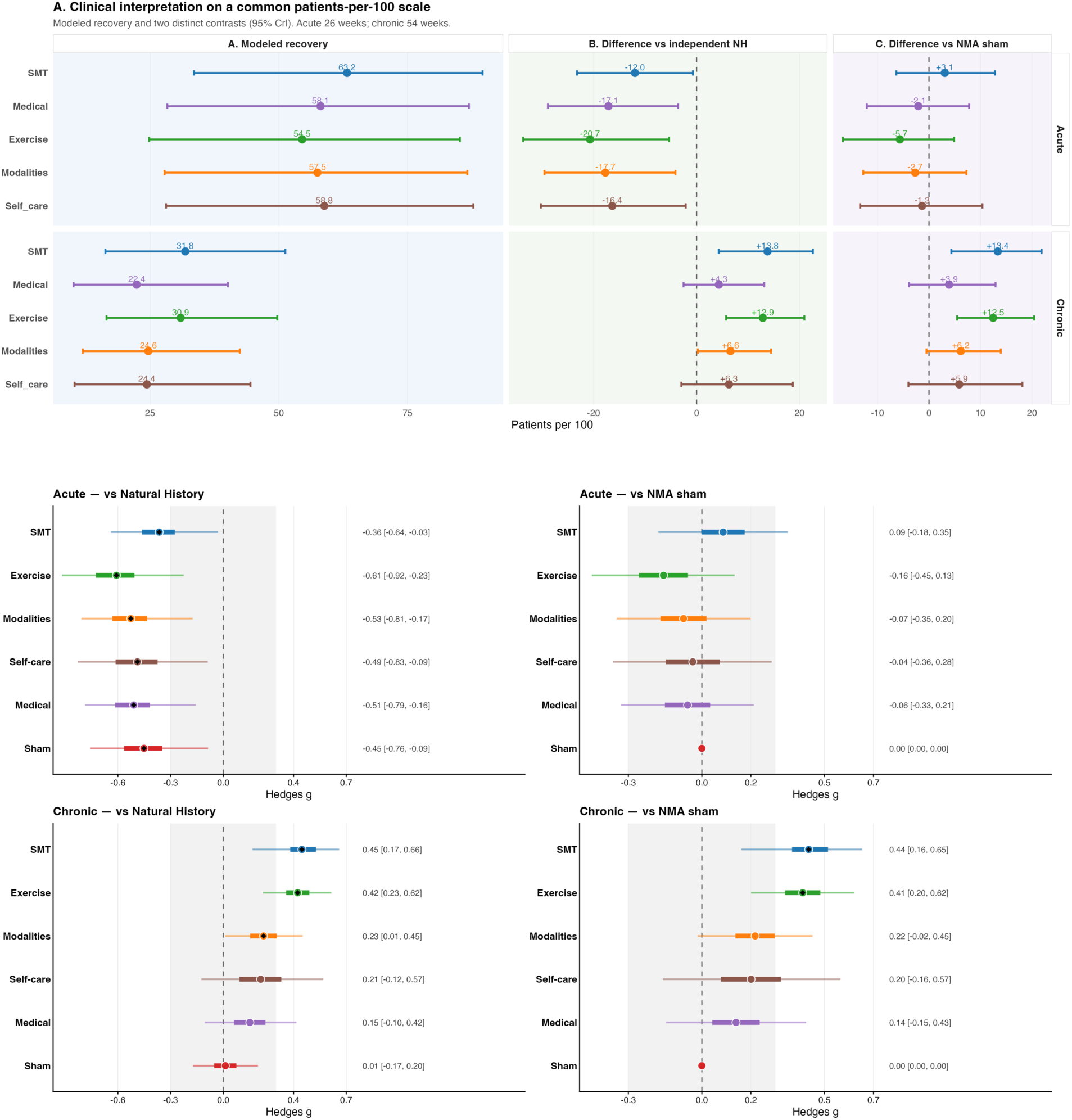
Clinical and standardized treatment effects. Acute and chronic results in clinical units are shown against the Paper I natural-history estimates. The standardized comparisons within the network can be read on their own, without the outside natural-history reference.

**Table 7.**
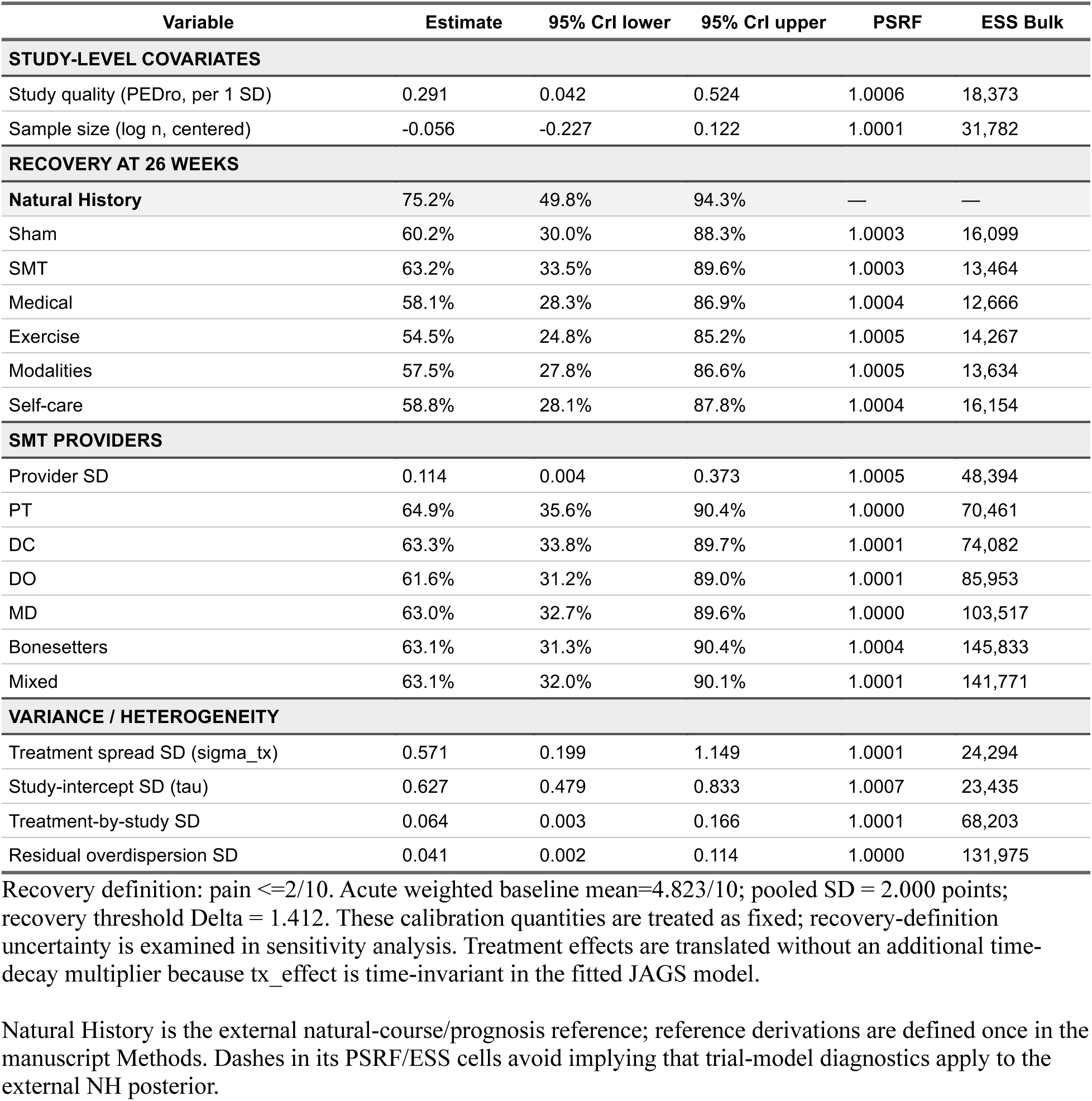
Bayesian model estimates for acute low back pain. Natural History is displayed with the trial treatments because it is the principal reference trajectory.

| Variable | Estimate | 95% CrI lower | 95% CrI upper | PSRF | ESS Bulk |
| --- | --- | --- | --- | --- | --- |
| **STUDY-LEVEL COVARIATES** |  |  |  |  |  |
| Study quality (PEDro, per 1 SD) | 0.291 | 0.042 | 0.524 | 1.0006 | 18,373 |
| Sample size (log n, centered) | -0.056 | -0.227 | 0.122 | 1.0001 | 31,782 |
| **RECOVERY AT 26 WEEKS** |  |  |  |  |  |
| **Natural History** | 75.2% | 49.8% | 94.3% | — | — |
| Sham | 60.2% | 30.0% | 88.3% | 1.0003 | 16,099 |
| SMT | 63.2% | 33.5% | 89.6% | 1.0003 | 13,464 |
| Medical | 58.1% | 28.3% | 86.9% | 1.0004 | 12,666 |
| Exercise | 54.5% | 24.8% | 85.2% | 1.0005 | 14,267 |
| Modalities | 57.5% | 27.8% | 86.6% | 1.0005 | 13,634 |
| Self-care | 58.8% | 28.1% | 87.8% | 1.0004 | 16,154 |
| **SMT PROVIDERS** |  |  |  |  |  |
| Provider SD | 0.114 | 0.004 | 0.373 | 1.0005 | 48,394 |
| PT | 64.9% | 35.6% | 90.4% | 1.0000 | 70,461 |
| DC | 63.3% | 33.8% | 89.7% | 1.0001 | 74,082 |
| DO | 61.6% | 31.2% | 89.0% | 1.0001 | 85,953 |
| MD | 63.0% | 32.7% | 89.6% | 1.0000 | 103,517 |
| Bonesetters | 63.1% | 31.3% | 90.4% | 1.0004 | 145,833 |
| Mixed | 63.1% | 32.0% | 90.1% | 1.0001 | 141,771 |
| **VARIANCE / HETEROGENEITY** |  |  |  |  |  |
| Treatment spread SD (sigma_tx) | 0.571 | 0.199 | 1.149 | 1.0001 | 24,294 |
| Study-intercept SD (tau) | 0.627 | 0.479 | 0.833 | 1.0007 | 23,435 |
| Treatment-by-study SD | 0.064 | 0.003 | 0.166 | 1.0001 | 68,203 |
| Residual overdispersion SD | 0.041 | 0.002 | 0.114 | 1.0000 | 131,975 |

Recovery definition: pain $\leq 2/10$ . Acute weighted baseline mean=4.823/10; pooled SD = 2.000 points; recovery threshold Delta = 1.412. These calibration quantities are treated as fixed; recovery-definition uncertainty is examined in sensitivity analysis. Treatment effects are translated without an additional time-decay multiplier because tx\_effect is time-invariant in the fitted JAGS model.
Natural History is the external natural-course/prognosis reference; reference derivations are defined once in the manuscript Methods. Dashes in its PSRF/ESS cells avoid implying that trial-model diagnostics apply to the external NH posterior.

Why are absolute recovery rates so uncertain while differences between treatments are much sharper? Because every treatment’s modeled recovery in Table 6 is built on the same natural-history draw: p_k(s) = Φ[*g*_NH(s) + θ_k(s) − Δ]. In a draw where natural history is favorable, all arms are high. In a draw where it is unfavorable, all are low. That shared uncertainty is why each absolute interval runs from roughly 25% to 90%. When we subtract one arm from another within the same draw, most of the shared part cancels out. What remains is uncertainty about the treatment difference itself. For example, the SMT-versus-sham interval spans about 19 points rather than 56. So wide absolute intervals and narrower differences do not conflict. They answer different questions.

In chronic LBP, modeled recovery (a favorable course) at 54 weeks was 18.1% for natural history (95% CrI 8.8% to 31.9%). It ranged from 18.5% for sham to 31.8% for SMT. Per 100 patients, SMT gave 13.8 more recoveries than natural history (95% interval +4.3 to +22.6), exercise 12.9 more (+5.7 to +20.9), and modalities 6.6 more (+0.2 to +14.5). The intervals for medical care (+4.3; −2.5 to +13.1) and self-care (+6.3; −3.0 to +18.7) included zero. Compared with sham, only SMT (+13.4; +4.3 to +21.9) and exercise (+12.5; +5.5 to +20.4) had intervals that excluded zero. Modalities (+6.2; −0.5 to +13.9), self-care (+5.9; −4.0 to +18.1), and medical care (+3.9; −3.9 to +12.9) were uncertain. No treatment reached 50% average recovery. Table 8 gives the full chronic model summary.

**Table 8.** Bayesian model estimates for chronic low back pain. Natural History is displayed with the trial treatments because it is the principal reference trajectory.

| Variable | Estimate | 95% CrI lower | 95% CrI upper | PSRF | ESS Bulk |
| --- | --- | --- | --- | --- | --- |
| **STUDY-LEVEL COVARIATES** |  |  |  |  |  |
| Study quality (PEDro, per 1 SD) | 0.052 | -0.119 | 0.219 | 1.0000 | 30,130 |
| Sample size (log n, centered) | -0.075 | -0.219 | 0.068 | 1.0001 | 35,743 |
| **RECOVERY AT 54 WEEKS** |  |  |  |  |  |
| **Natural History** | 18.1% | 8.8% | 31.9% | — | — |
| Sham | 18.5% | 8.4% | 33.6% | 1.0001 | 52,643 |
| SMT | 31.8% | 16.3% | 51.3% | 1.0004 | 18,588 |
| Medical | 22.4% | 10.1% | 40.1% | 1.0000 | 70,327 |
| Exercise | 30.9% | 16.5% | 49.7% | 1.0001 | 44,037 |
| Modalities | 24.6% | 11.9% | 42.4% | 1.0001 | 57,306 |
| Self-care | 24.4% | 10.3% | 44.5% | 1.0001 | 76,156 |
| **SMT PROVIDERS** |  |  |  |  |  |
| Provider SD | 0.143 | 0.006 | 0.459 | 1.0026 | 17,555 |
| PT | 35.5% | 19.9% | 55.1% | 1.0007 | 23,453 |
| DC | 32.6% | 17.7% | 51.7% | 1.0005 | 25,616 |
| DO | 33.2% | 17.8% | 53.0% | 1.0006 | 28,440 |
| MD | 29.8% | 14.6% | 49.2% | 1.0002 | 49,384 |
| Bonesetters | 32.9% | 16.7% | 53.6% | 1.0001 | 40,560 |
| Mixed | 32.1% | 13.0% | 55.0% | 1.0017 | 160,809 |
| **VARIANCE / HETEROGENEITY** |  |  |  |  |  |
| Treatment spread SD (sigma_tx) | 0.361 | 0.166 | 0.722 | 1.0005 | 57,144 |
| Study-intercept SD (tau) | 0.433 | 0.331 | 0.558 | 1.0001 | 82,300 |
| Treatment-by-study SD | 0.115 | 0.009 | 0.229 | 1.0002 | 35,285 |
| Residual overdispersion SD | 0.035 | 0.001 | 0.099 | 1.0000 | 131,640 |

Recovery definition: favorable natural course at 54 weeks, the reference (DIRECT) outcome family of the 12-cohort chronic Natural History model. Treatment recovery = $\Phi(\Phi^{-1}(p_{\text{NH}}) + \text{effect}_k)$ , using independently paired NH and treatment-model draws; the chronic treatment model was fitted with this NH posterior as its external offset. Chronic NH convergence: max PSRF 1.0029, min ESS 3,226.
Natural History is the external natural-course/prognosis reference; reference derivations are defined once in the manuscript Methods. Dashes in its PSRF/ESS cells avoid implying that trial-model diagnostics apply to the external NH posterior.

**Table 9.** Acute treatment contrasts under primary saturating-NH and Bayesian linear-NH.

| Treatment | Primary | Bayesian linear NH | P(better than sham),<br>linear NH |
| --- | --- | --- | --- |
| SMT | 0.087 [-0.177, 0.351] | 0.083 [-0.179, 0.346] | 0.742 |
| Medical | -0.058 [-0.330, 0.212] | -0.064 [-0.335, 0.206] | 0.320 |
| Exercise | -0.156 [-0.449, 0.134] | -0.150 [-0.438, 0.137] | 0.154 |
| Modalities | -0.075 [-0.348, 0.198] | -0.089 [-0.361, 0.183] | 0.260 |
| Self-care | -0.037 [-0.363, 0.285] | -0.055 [-0.376, 0.263] | 0.371 |

Values are posterior mean standardized treatment contrasts versus NMA sham with 95% credible intervals. The Bayesian linear-NH model is a deliberately stringent functional-form sensitivity analysis; the primary saturating NH model remains authoritative.

Treatment effects were smaller against sham than against natural history. In chronic LBP, sham and natural history were nearly identical (+0.4 recoveries per 100; 95% interval −4.2 to +5.7). We read this difference as placebo responsiveness: the extra improvement from receiving sham, after removing the recovery expected anyway. It reflects the patient’s response, not a property of the sham itself. Here it was close to zero. SMT and exercise showed the clearest effects beyond sham. But the active treatments overlapped a great deal, so the data do not support a firm ranking.

### Sensitivity to the definition of clinically meaningful recovery

Figure 5 asks whether the advantage of each treatment depends on how we define a meaningful outcome. For each active treatment, it plots the difference from sham in recovery probability under four definitions: any improvement, at least 30% reduction, at least 50% reduction, and pain ≤2/10. In acute LBP, treatment-versus-sham differences stayed close to zero under every definition. In chronic LBP, SMT and exercise stayed above sham under all four definitions, with every 95% interval above zero. Medical care, modalities, and self-care had intervals that included zero under all four. For chronic LBP, the alternative definitions were applied as threshold shifts relative to the favorable-course anchor, so they are approximate. Figure 5 thus tests how robust the treatment-versus-sham comparison is. It does not test absolute recovery.

**Figure 5.**
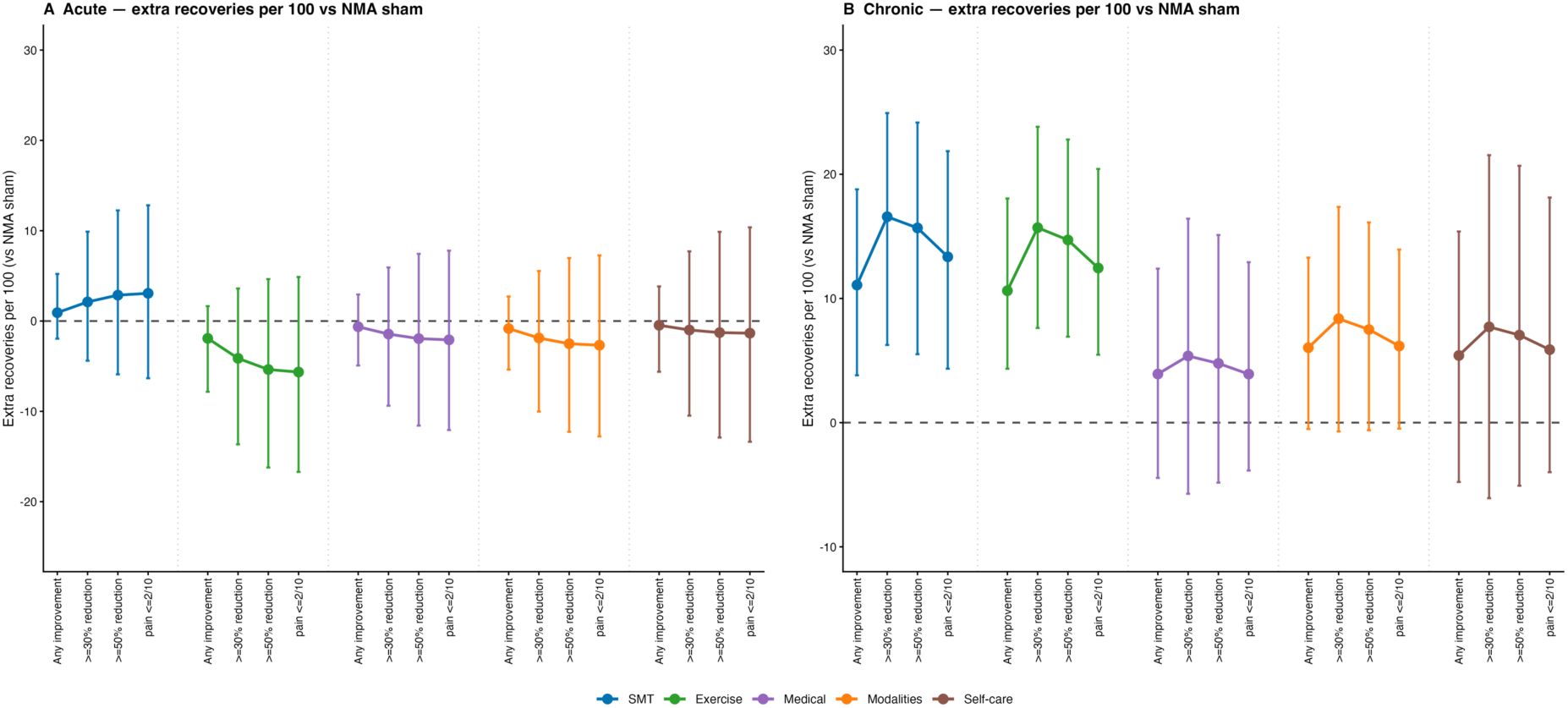
Sensitivity of treatment advantage to the definition of a meaningful outcome. For each active treatment, points show the posterior mean number of extra recoveries per 100 compared with sham, under four recovery definitions. Bars are 95% credible intervals. Acute estimates are at 26 weeks and chronic estimates at 54 weeks. The zero line means no advantage over sham. Sham is the reference and is not plotted.

Across all four definitions, the active treatments stayed close together in acute LBP. This supports reading the results separately by phase. It also warns against ranking treatments by posterior means alone.

Together, Figures 3–5 separate overall prognosis from the advantage of one treatment over another. Figure 3 shows absolute recovery over time for natural history and each treatment.

Figure 4 shows modeled recovery along with differences from natural history and from sham. Figure 5 asks whether the treatment-minus-sham difference holds up under other definitions of a meaningful outcome. In acute LBP, background recovery is strong and treatment adds little. In chronic LBP, the outlook is worse overall, but SMT and exercise show clearer added benefit over sham.

## Natural history, contextual effects, and treatment-specific effects

The pattern of comparisons differed by phase. In acute LBP, natural history was more favorable than the treatment arms at 26 weeks. Sham fell below natural history (−15.0 per 100; −27.7 to −2.1). Treatment-versus-sham differences were close to zero. In chronic LBP, active treatments were above natural history. Sham was essentially equal to it (+0.4 per 100; −4.2 to +5.7). This pattern lets us separate background prognosis from the added effect of treatment. It also lets us read sham minus natural history as placebo responsiveness, after removing the recovery expected anyway.

This placebo response may reflect expectation, the patient–provider relationship, reassurance, attention, conditioning, the ritual of treatment, provider interaction, and other effects of receiving a sham or ineffective treatment in a clinical or research setting. This analysis does not separate these mechanisms.

The overall pattern is best stated with care. In acute LBP, improvement happens against a very favorable natural course, and treatment adds little beyond sham. In chronic LBP, improvement happens against a less favorable natural course, and SMT and exercise show the clearest added effects beyond sham. Large improvements within treatment arms and modest differences between treatments can therefore exist side by side in the same evidence.

## Study quality and blinding

In acute LBP, studies with higher PEDro scores showed more improvement across all their arms (0.291 per SD; 95% CrI 0.042 to 0.524). In chronic LBP, the PEDro term was small and uncertain (0.052; −0.119 to 0.219). Because this term shifts each study’s overall level, it does not change comparisons between treatments within a study. Exploratory analyses of quality and blinding did not find a stable link between study quality and treatment differences. Estimates for different SMT provider types were also imprecise. Every 95% credible interval included zero in both acute and chronic LBP. So we found no consistent differences in SMT results by provider type. But overlapping intervals do not prove that providers are equivalent. Variation between providers was smaller than variation between studies (acute provider SD 0.114 versus τ 0.627; chronic 0.143 versus 0.433). Treatment-by-study and residual variation were smaller still.

The six acute provider estimates in Table 7 range only from 61.6% to 64.9%. That narrow range is partly a product of partial pooling. The model estimates how much providers truly differ (provider SD 0.114, interval from near zero to 0.373). It then pulls each provider’s estimate toward the SMT average. The less data a provider has, the stronger the pull. When the estimated spread is small and provider data are sparse, the estimates end up close together. So nearly identical provider estimates mean the data cannot tell providers apart. They do not show that providers are the same [181].

## Transitivity, exchangeability, and model assumptions

We analyzed the acute and chronic networks separately. Their background recovery differs too much to assume the two populations are exchangeable. Within each phase, measured effect modifiers overlapped enough to support exchangeability, but only conditionally. Direct and indirect estimates generally agreed. The main local inconsistency was modalities versus SMT in the acute network (P = 0.039). We found no local inconsistency in the chronic network. We treated sparse comparisons and leftover heterogeneity as reasons for caution, not as grounds to assume the studies were fully exchangeable.

No single measured covariate showed a broad failure of transitivity. Leftover differences between studies were handled by study-level random effects. We read the network estimates as average effects across trials that differ but overlap enough to combine. The direct–indirect comparisons in Figure 2 give an empirical check on consistency. The audit of clinical and design features supports exchangeability only conditionally.

## Sensitivity to the imported natural-history posterior

Under the linear-form acute natural-history posterior from Paper I, natural recovery at 26 weeks was 79.9% (95% CrI 53.8% to 95.7%), compared with 75.2% under the main saturating model. The active-treatment comparisons with sham hardly moved. SMT versus sham was *g* = 0.083 (95% CrI −0.179 to 0.346) under the linear model. Under the main saturating model, it was *g* = 0.087 (95% CrI −0.177 to 0.351). All other treatment-versus-sham intervals included zero under both models (Table 9; Figure 6).

**Figure 6.**
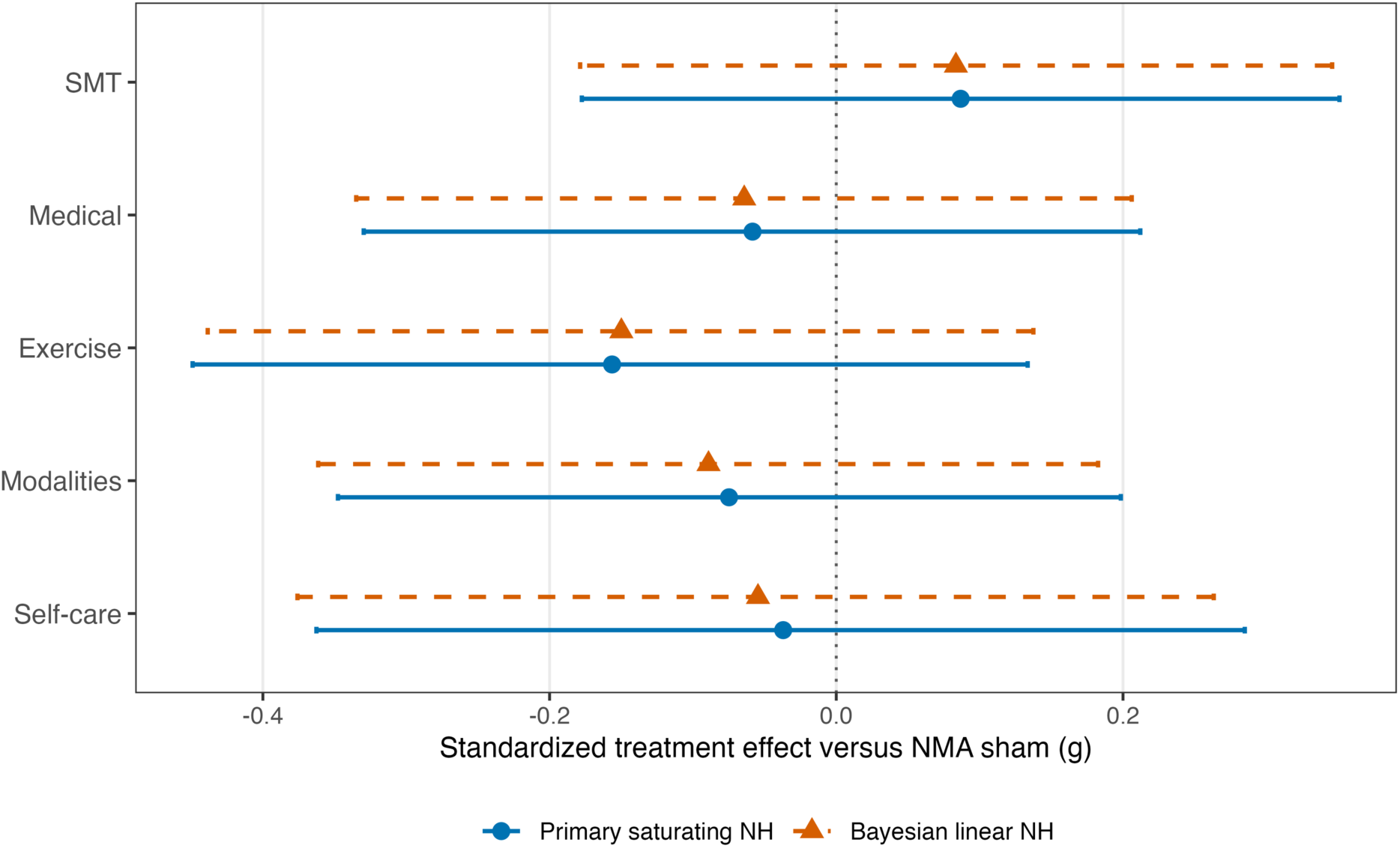
Sensitivity of acute treatment effects to the natural-history functional form. Standardized treatment effects versus NMA sham (g; posterior mean and 95% credible interval) under the primary saturating natural-history trajectory and under the Bayesian linear trajectory (Table 9).

In the acute common-asymptote sensitivity model (233 observations from 55 studies, fitted with lme4), treatments could speed up or slow down recovery but had to share the natural-history start and endpoint. Sham and every active treatment except self-care recovered more slowly than natural history. Rate shifts versus natural history were: sham −0.47 (95% CI −0.63 to −0.30), SMT −0.39 (−0.52 to −0.25), exercise −0.51 (−0.72 to −0.30), medical care −0.70 (−0.96 to −0.45), modalities −0.46 (−0.63 to −0.28), and self-care −0.34 (−0.82 to 0.15). Modeled recovery at 4 weeks was 63.9% for natural history against 22.9% to 43.2% across trial arms. By 12 weeks the gap had narrowed to under 5 points (natural history 76.7%; trial arms 72.4% to 74.7%), and at 26 weeks modeled recovery was 77.1% in every trial arm and 77.2% for natural history.

We also fitted a free-ceiling version of this model, in which each arm could differ from natural history both in its speed of recovery and in the level at which it leveled off. Letting the ceiling vary fitted the data far better than forcing convergence (AIC 319.8 versus 465.4; likelihood-ratio χ² = 157.6, 6 df, P < 0.001), and every arm’s ceiling lay below natural history (for example, SMT −1.05 *g*, 95% CI −1.21 to −0.89). Over the observed follow-up, then, the gap between trial arms and natural history did not close. The size of that gap, however, depended on the model. The free-ceiling model, which lacks the primary model’s treatment-by-study random effects and holds natural history at its posterior-mean curve, gave much larger gaps and implausible shapes, with sham recovery rising to 27.2% at 4 weeks and then falling. We therefore take the primary model’s within-trial estimates as the more conservative measure of the gap, and draw from the time-shape analyses only the conclusions they share: trial arms recover more slowly than natural history early, and the gap persists through the observed follow-up.

These results show that absolute recovery is sensitive to the natural-history model, but comparisons between treatments are not. Acute sensitivity to curve shape is reported here, using the linked Paper I posterior. Chronic natural-history sensitivity, validation against sources, and decisions about outcome types belong to Paper I. This paper reports sensitivity within the treatment network and what happens when other approved Paper I posteriors are imported.

Other recovery definitions, treatment-time translations, acute node-splitting time points, scale reconciliation, and common-asymptote checks did not change the main conclusions. Acute LBP showed fast background recovery and little treatment-versus-sham difference. Chronic LBP showed slower natural recovery, with SMT and exercise giving the clearest added benefit over sham. Figure 5 shows that the acute treatment-versus-sham conclusions hold under all four recovery definitions.

## Discussion

### Principal Findings

This analysis helps explain a long-standing puzzle in the low back pain (LBP) literature. Patients usually get better, yet it has been hard to show which treatment is responsible. Much of the puzzle goes away once three quantities are kept apart. They are often blurred together. The first is absolute recovery: how many patients get better. The second is improvement beyond the expected natural course. The third is improvement beyond sham.

This is a counterfactual analysis, not a ranking of treatments. The question is not whether patients improved but compared with what. Natural history and treatment were each estimated as full probability distributions. So the result compares two distributions, rather than subtracting one point estimate from another [38].

The answer depended strongly on how long the pain had lasted. In acute LBP, natural history showed fast improvement. Recovery rates on treatment were high. But once this background recovery was considered, little extra improvement was left for treatment. Effects relative to natural history were mostly negative or near zero. Effects relative to sham were near zero, with wide uncertainty. Acute LBP thus shows that a good outcome does not mean a large treatment effect. Many patients recover while being treated, even when little of that recovery is due to the treatment.

The acute shortfall against natural history should not be read as proof that treatment slows recovery. No treatment, not even sham, recovered faster than natural history. On average, treated arms were somewhat slower (Table 6). The common-asymptote rate model points the same way. Three features of the evidence argue against a causal reading. First, sham also fell below natural history, and no active treatment beat sham. So, the shortfall is shared by all trial arms and cannot be pinned on any one treatment. Second, the comparison is between trial arms and outside cohorts, not a randomized comparison. Trial patients are care-seekers selected for pain severity and duration, and outcome definitions differ between trials and cohorts. Third, the size of the shortfall depends on which curve shape is assumed for acute natural history (Table 9). A true causal effect of treatment would not. These data cannot tell whether the shortfall comes from trial participation, the selection of care-seekers, or treatment itself (for example, by making patients feel more ill, or through nocebo effects).

Chronic LBP showed the opposite pattern. Natural history predicted much slower and less complete recovery (18.1% at 54 weeks). Absolute recovery was therefore lower than in acute LBP. SMT, exercise, and modalities beat natural history, and SMT and exercise showed the clearest benefit over sham. Placebo responsiveness (sham minus natural history) was close to zero (+0.4 per 100; −4.2 to +5.7). In chronic LBP, then, receiving sham added little beyond the expected course. Absolute prognosis and the comparative effect of treatment remain two different things.

Why has the SMT literature stayed so mixed? Our findings suggest that the answer differs between acute and chronic LBP. In acute LBP, fast natural improvement creates a large background signal. Small treatment-specific differences must be detected against it. So, a large improvement in an SMT arm is not, by itself, evidence that SMT works. Repeated trials may keep producing favorable, null, or unfavorable results, depending on chance, the comparator, and the follow-up time. In chronic LBP, natural recovery is weaker, so a treatment signal is easier to see. Here, SMT and exercise showed the clearest advantage over sham. This acute–chronic difference may explain why the SMT literature remains inconclusive. It also closely matches an earlier comparative-effectiveness analysis. In that study, nonspecific factors explained most acute improvement, while treatment explained a larger share of chronic outcomes [182]. Even in chronic LBP, though, modeled recovery at 54 weeks stayed below 50% (SMT 31.8%; exercise 30.9%), with wide uncertainty.

### Natural history and the Interpretation of Treatment Response

Most treatment studies assume natural history rather than estimate it. Randomization protects the comparison between groups from many biases. But it does not show that the improvement within either group was caused by the treatment. When a condition improves quickly on its own, a treatment can look very successful in a before-and-after or within-group analysis, even when its added effect is small [9, 183].

The central finding is the difference between doing better than a comparator and reaching an adequate level of recovery. For uncomplicated acute LBP, the natural-history evidence shows a mostly self-limiting course. Much of the recovery happens in the first several weeks, with no clear contribution from treatment. The treatment analyses give little evidence that any of the treatments speed this recovery. Every treatment, sham included, fell below natural history, with intervals that excluded zero. As discussed above, this more likely reflects differences between trial patients and natural-history cohorts than harm from treatment. In acute LBP, the key clinical question may not be whether patients improve with treatment. It may be whether treatment speeds recovery beyond what would happen anyway. Here, there was little evidence that it did. In chronic LBP, SMT and exercise show the clearest added benefit over sham. Yet their average recovery remains below the 50% benchmark (SMT 31.8%; exercise 30.9%). That benchmark describes the population. It is not the chance that any one patient will benefit from treatment.

A descriptive sensitivity analysis of studies that enrolled mixed acute and chronic patients did not show results falling consistently between the acute and chronic estimates. This supports assigning studies to a phase, rather than treating mixed enrollment as a separate phase.

This analysis tackles the problem directly. It uses a natural-history estimate built separately from the treatment network (Paper I). It treats that estimate as uncertain rather than fixed. That uncertainty is then carried through the treatment analysis.

Splitting acute from chronic LBP is not just a convenience. It reflects two different background processes, which are described in Paper I.

### Placebo Responsiveness

Sham meant a placebo procedure or a treatment known to be ineffective. We separate the sham procedure from the placebo response, which is the patient’s response to receiving it.

Improvement seen with sham has two parts: the natural course of LBP, and the response to the sham. We therefore estimated placebo responsiveness as sham minus natural history. By removing the improvement predicted from independent natural-history evidence, this gives a cleaner estimate of the placebo response than raw improvement on sham.

A simple example makes the point. Suppose 68 of 100 sham-treated patients recover. Suppose natural history predicts that 60 of 100 similar patients would have recovered anyway (hypothetical values). Most of the sham result then reflects the natural course. Only the difference, 8 per 100, is due to receiving sham in a clinical setting. Calling 68 per 100 the placebo effect would overstate it more than eightfold. This is the difference between improvement in a placebo group and the placebo effect itself [9, 183].

Placebo responsiveness happens within the patient. It is not a property of the sham. Expectation, the patient–provider relationship, reassurance, attention, conditioning, the ritual of treatment, and provider interaction may all play a part. This analysis does not try to separate them. More broadly, placebo responsiveness has been proposed as an evolved capacity. In this view, cues from the environment and from other people can change the timing or size of the body’s own responses to illness, pain, and recovery. Humphrey suggested that this responsiveness may help the body manage costly physical resources. Trimmer and colleagues later formalized this idea in evolutionary models [184–187].

This analysis does not test that evolutionary idea. Instead, by separating natural-history improvement from the extra response seen with sham, it gives an empirical estimate of placebo responsiveness. It does not say how that responsiveness arises.

### Extensive Overlap Among Conservative Treatments

A key feature of this analysis is how much the main treatment categories overlap once uncertainty is considered. The absolute recovery estimates and the relative effects tell the same story. SMT and exercise tend to sit near the top of the chronic group. But the gaps among active treatments are small compared with the uncertainty and the differences between studies.

Point estimates and rankings can make small differences look bigger than they are [188]. In acute LBP, the key feature is that treatment-versus-sham effects cluster tightly around zero, against a very favorable natural course. In chronic LBP, SMT and exercise show the clearest effects beyond sham. But the other active treatments overlap widely, and even the top two overlap each other. A consistent numerical order is not evidence that one treatment is meaningfully better.

The evidence supports a firmer conclusion about the difference between prognosis and treatment effect than about the exact order of treatments. In chronic LBP, SMT and exercise are the only active treatments with clearly positive effects versus sham in the current run. The evidence is less clear for medical care, modalities, and self-care. This is a narrower claim than saying that all structured conservative care reliably beats natural history or sham.

### Between-Study Heterogeneity

Differences between studies remained large. This is not surprising, given how varied the LBP literature is. Studies differ in which patients they enroll, how severe and how long-lasting their pain is, how much treatment they give, how providers behave, how the comparator is designed, when outcomes are measured, how well the study is done, and how outcomes are measured.

This variation is informative. When differences between studies are large compared with differences between treatments, trials can honestly reach different answers. One does not have to be “right” and the other “wrong.” Each is observing the treatment effect in a different clinical and research setting.

Standardized numbers are easier to judge when turned back into pain points. In our setup, 1 *g* equals about 2.0 NRS points. In acute LBP, the SD of study-level improvement was τ = 0.627 *g*, so about 95% of studies’ underlying improvement lies within roughly two τ of the average. By comparison, the acute SMT-versus-sham effect was *g* = 0.09. The spread between studies is thus many times larger than the treatment difference we are trying to detect. That is why individual trials can disagree without any of them being wrong [189].

Standard heterogeneity statistics show that variation exists, but not where it comes from. The hierarchical model used here separates study-level variation from treatment effects. At the same time, it takes the expected natural course into account. The results suggest that much of the apparent disagreement about SMT may instead reflect differences in study populations, timing, sham conditions, and other design features.

### Network Assumptions and Consistency

Overall, the network checks supported interpreting the NMA estimates within each phase. Exchangeability should be seen as conditional, not absolute. Measured effect modifiers overlapped enough to combine studies. But leftover heterogeneity and thin evidence for some comparisons call for caution. Direct and indirect estimates generally agreed. Where they disagreed or evidence was thin, we interpreted the network estimates more cautiously. The network estimates are best read as average effects across varied but reasonably comparable studies. They are not effects that will apply everywhere.

Absence of evidence is not evidence of absence. Two conclusions in this paper rest on this logic. A provider estimate whose interval includes zero does not show that providers are equivalent. A node split that finds no inconsistency does not show that direct and indirect evidence agree. In both cases, the interval may simply be too wide to reveal a real difference. Equivalence can be claimed only when the whole credible interval falls inside a preset zone of no practical importance, such as the ±0.30 zone used here [170, 190].

### Study Quality and Blinding

Study quality may still help explain why LBP studies differ. Clinicians who deliver hands-on treatment usually cannot be blinded, and convincing patient blinding can be hard. In our models, higher-quality acute studies showed more improvement across all arms, while the chronic PEDro term was small and uncertain. Because the term shifts whole studies, it does not by itself explain differences between treatments.

PEDro is a combined quality score, not a pure measure of blinding. So these results cannot show which design features, if any, affect treatment effects. Because treating clinicians cannot be blinded, the provider’s interaction with the patient is bound up with many hands-on treatments. This makes credible sham procedures, matched for attention, especially valuable when the question is the specific effect of manipulation.

### Clinical Interpretation

The evidence is clearest for acute LBP. No treatment studied, SMT included, added a meaningful benefit over sham, and no treatment arm recovered more often than natural history. For uncomplicated acute LBP, the evidence therefore supports watchful waiting: reassurance, advice to stay active, and a supportive clinical relationship, with reassessment if recovery stalls or warning signs appear. Because LBP is among the costliest conditions in health care, routinely front-loading treatment in acute LBP spends resources where natural recovery is already strong [2]. Chronic LBP is different. There, SMT added about 13 recoveries per 100 patients at 54 weeks beyond both sham (+13.4; 95% interval +4.3 to +21.9) and natural history (+13.8; +4.3 to +22.6), and exercise added a similar amount over sham (+12.5; +5.5 to +20.4).

In chronic LBP, natural improvement is slower and overall recovery is lower. Yet SMT and exercise show clearer added benefit over sham. The clinical question therefore shifts. It is no longer whether patients improve, but how much extra benefit comes from the chosen treatment. The sham experience may still differ across trials.

In uncomplicated acute LBP, fast expected recovery should be central to how results are read. Our results support reassuring patients about their outlook and managing them conservatively. They give little overall evidence that any of the treatments studied changes the underlying recovery path. This does not mean treatment can never relieve symptoms or suit an individual patient. It means that improvement after treatment should not, by itself, be taken as proof that treatment caused or sped up recovery [16, 191, 192].

Because active treatments overlap so much, choosing a treatment for chronic LBP can reasonably weigh safety, cost, access, patient preference, and burden. It can also weigh how well a treatment encourages activity and self-management. Small differences in average effectiveness need not be the only guide.

#### The cost and risk of needless clinical care

Treatment costs do not end with the fee. The average US trip for medical care is about 10 road miles each way[193], and an ambulatory visit takes about 121 minutes, 37 of them traveling and 84 in the clinic [194]. A course of 12 visits therefore adds about 240 miles and 24 hours. Vehicle operating costs at the IRS medical mileage rate of $0.235 per mile add about $56 [195]. External costs of driving, including congestion, local air pollution, and crash risk, add about $39 at $0.163 per mile [196] (in 2026 dollars).

Patient time, valued at about $66 per visit [194](in 2026 dollars), adds about $797. Together these come to roughly $875–$965 per course, depending on whether the patient lives in an urban or a rural area, before any fee is paid. Adding the mean amount paid per chiropractic visit in national survey data, about $114 in 2026 dollars ($84.84 in 2018 dollars) [197], brings the total to roughly $2,240–$2,330 per course.

Treatment also carries iatrogenic risk. Minor, transient adverse events, such as soreness or increased pain, follow spinal manipulation in about half of patients [198]. Serious events after lumbopelvic manipulation, including cauda equina syndrome, disc herniation, and fracture, are known only from case reports, so their incidence cannot be estimated, although it is thought to be very low [199]. Every comparator carries its own risks; neither medical care nor sham is risk-free.

At the population level, the Medical Expenditure Panel Survey records about 31–44 million chiropractic visits each year by US adults linked to low back conditions [200]. Because many chiropractic visits carry no coded musculoskeletal diagnosis, the true number is probably higher. Fees, travel, externalities, and patient time for these visits total about $6–8.5 billion annually (2026 dollars). In acute LBP, where no treatment outperformed natural history, this spending buys no recovery beyond what natural history already provides. In chronic LBP, SMT’s added benefit over sham of about 13 recoveries per 100 patients implies roughly $17,000 per additional recovery, counting all costs (about $10,200–$54,200 across the 95% interval).

A main strength of this study is that it keeps natural-history evidence separate from the treatment network. Rather than assuming a fixed natural course, we imported an independent natural-history estimate (Paper I). We then carried its uncertainty into the treatment comparisons.

A second strength is that we report both absolute and comparative results. Treatment versus natural history asks whether improvement exceeds the expected course of the condition.

Treatment versus sham asks whether a treatment adds benefit beyond the sham experience. Recovery probabilities describe how many patients are modeled to reach a meaningful outcome. These are different questions. Looking at them together guards against two errors. A high recovery rate is not mistaken for a large treatment effect. And a real added effect in chronic LBP is not dismissed just because many patients still do not fully recover.

Other strengths include separate acute and chronic analyses, explicit study-level random effects, checks of transitivity and exchangeability, sensitivity analyses of recovery definitions and natural-history models, assessment of study quality, and an audit that kept the natural-history evidence independent of the treatment network. We also displayed full distributions, so that overlap between treatments could be judged directly rather than from point estimates or rankings alone.

## Limitations

This study has several important limitations. First, the imported natural-history estimate shares the limitations of its evidence base, which are described in Paper I.

Second, natural-history cohorts, randomized trial populations, and observational treatment cohorts were not randomized against each other. So they cannot be assumed to be fully comparable. Observational cohorts were included only when their population, outcomes, timing, clinical phase, and source supported the role they played. Study-level random effects and measured study features reduce leftover differences but cannot remove them. Observational cohorts were never used in place of randomized treatment comparisons.

Third, sham procedures and trial settings varied across studies, even though sham was limited to placebo procedures or treatments known to be ineffective. This variation limits what the sham-minus-natural-history placebo estimate can tell us about mechanisms.

Fourth, putting different pain scales on a common footing requires assumptions. The conversion to recovery probability is easy to interpret clinically. But it depends on the chosen threshold and on assumptions about how individual changes are distributed. We used sample-size-weighted baseline means for each phase (4.823 acute; 5.284 chronic) and a fixed SD_NRS of 2.0. These gave thresholds of Δ = 1.412 and 1.642. We treated these values as fixed, not as uncertain.

Testing other recovery definitions reduces, but cannot remove, our reliance on the chosen threshold and conversion. In chronic LBP, the alternative recovery definitions were applied as threshold shifts relative to the favorable-course anchor, so those sensitivity results are approximate.

Fifth, the treatment trials mostly enrolled patients who could walk and attend outpatient care. Patients with very severe pain, major loss of function, inability to walk, worsening nerve damage, or other conditions that rule out outpatient trials may be under-represented. These estimates should be applied cautiously to more severe or non-ambulatory patients.

Sixth, study quality was rated once per study, with AI assistance, and the ratings were not validated against human reviewers.

Seventh, treatment effects were assumed constant over time. Trial follow-up was short relative to the 26-and 54-week horizons, so absolute comparisons with natural history at those horizons are extrapolations. The common-asymptote and free-ceiling sensitivity analyses test other time shapes.

Finally, this is a study-level synthesis. Patient characteristics that may change response cannot be properly studied without individual participant data. Individual participant data meta-analyses of SMT in chronic LBP have begun to address this [201, 202]. Overlap in average treatment effects does not mean that all patients respond the same way. Nor does it rule out useful subgroups of patients who respond differently.

## Implications for Future Research

Future LBP trials could be far more informative if they were designed around three things: natural recovery, the general effects of receiving care, and the specific effects of treatment.

In acute LBP, researchers should remember that treatment is being judged against a fast-changing background. Outcome times should be set in advance with expected natural recovery in mind. Claims that a treatment speeds recovery should require designs that can separate its effect from that background.

When the aim is to test a specific treatment mechanism, sham conditions should match provider contact, expectation, visit frequency, and other features of care as closely as possible. Comparing a hands-on treatment with an untreated waitlist cannot tell whether any difference comes from the procedure or from the clinical visit.

Future natural-history studies would also benefit from standard pain measures and repeated measurements starting close to symptom onset. They should record any treatment patients receive during follow-up. They should report enough data to estimate both the average course and its uncertainty. Individual participant data would allow more direct modeling of how recovery varies, and of possible subgroups that respond differently.

Finally, comparative-effectiveness studies should report uncertainty and overlap, not just treatment rankings. When several treatments have largely overlapping results, it may be more useful to find the safest, least burdensome, and most acceptable way to deliver the helpful parts of care they share. That may matter more than chasing small differences in average effect.

The clinical decision is not which treatment ranks first. It is whether the leading treatment helps enough to justify its use. In chronic LBP, SMT and exercise show added benefit over sham. Even so, their average recovery stays below 50% at 54 weeks. The analysis does not identify the individual practitioner as the active ingredient. Practitioner, patient, technique, and their interactions all remain possible sources of variation. Sorting them out will require linked data and replication.

## Conclusion

This analysis separates the large improvement seen during treatment from the added benefit of treatment over a comparator. In acute LBP, natural recovery was favorable, and treatment showed little reliable advantage over sham. In chronic LBP, SMT and exercise showed the clearest added effects over sham. Even so, their modeled recovery stayed below 50% (31.8% and 30.9%). These findings call for a more cautious view of what treatment specifically achieves.

Care that adds nothing to natural recovery is not free: in acute LBP, travel, patient time, fees, and iatrogenic risk are incurred without additional benefit, at roughly $2,240–$2,330 per 12-visit course. These costs are part of the roughly $6–8.5 billion spent each year on US chiropractic visits for low back conditions. Differences between individual patients and between practitioners remain unresolved.

These findings may explain a long-standing feature of the SMT literature. Repeated trials have produced estimates that are often small, inconsistent, and frustratingly unstable. The treatment signal must be found against large natural improvement, placebo responsiveness, and differences between studies. When improvement in the comparison arm is high, the sample needed to detect a fixed treatment advantage grows steeply: with 59% improving on active treatment, about 72 participants are needed if 24% improve on placebo, but about 366 if 44% do [203]. Against that background, small differences in patients, comparators, follow-up, and design can change the observed result. Repeating the same kind of study therefore does not necessarily settle the question. More unstable estimates can make the literature bigger without making the treatment effect clearer. Future SMT research may be more useful if it is designed to separate treatment-specific benefit from natural history and the general response to care. Simply adding more conventional comparisons to an already large evidence base is unlikely to help.

“Treatment works” is an incomplete scientific statement unless we say compared with what [38]. Estimating the expected course independently does not replace randomized evidence. It changes the question we can ask of it. The question moves from “Did patients improve?” to “How much better did patients do than we would otherwise have expected?”

Future research should focus on questions that could change clinical decisions. Formal value-of-information analysis could test whether reducing the remaining uncertainty is worth more trials.

Studies that link patients to practitioners could look for consistent differences in how patients respond.

## From the “tic” to the “tor”

Chiropractic has long distinguished between the “’tic,” meaning the profession or system of care, and the “’tor,” meaning the individual chiropractor. Putting the profession first implies a clinical product that can be transferred from one practitioner to another. If so, its benefits should be reproducible across practitioners.

This pooled evidence cannot show whether the practitioner, the technique, patient characteristics, or their interactions explain the leftover variation. Answering that will need data that link patients to practitioners, with enough replication to estimate each source of variation. An explanation based on the individual practitioner remains possible, but it has not been tested.

Under our model, outcome definition, and clinical benchmark, the evidence does not reliably show a general, clinically meaningful benefit specific to SMT as a category. This does not show that all effects of manipulation are zero. Nor does it show that individual practitioners cannot achieve good results, or that a professional title marks an effective ingredient. The roles of patient, practitioner, technique, and context remain unresolved.

The value of more conventional trials should be judged by the decisions they could change. Formal value-of-information analysis could weigh the expected benefit of reducing uncertainty against research costs. Without such an analysis, this study cannot measure the cost or futility of further research.

## Declarations

### Ethics approval and consent to participate

Not applicable. This study synthesized published aggregate data and involved no new data collection from human participants.

### Consent for publication

Not applicable.

### Availability of data and materials

The arm-level dataset is available at Zenodo (doi:10.5281/zenodo.23609810). The R and JAGS code for all models, tables, and figures, the saved model outputs, and the data dictionary are available at Zenodo (doi:10.5281/zenodo.23655011). The natural-history evidence and models are described in the companion paper (Paper I).

### Authors’ contributions

JMM: concept, writing, and analysis. AJF: concept, writing, and analysis. MP-A: writing, manuscript review, and analytic review. All authors read and approved the final manuscript.

## Data Availability

Updated data and analytic materials underlying the results are available from the Open Science Framework (OSF) repository: https://osf.io/y5uh4/

## Acknowledgements

We thank Dan Malone, PhD, and Lauren E. Menke, BS (MD student).

## Abbreviations

AI: Akaike information criterion
CI: confidence interval
CrI: credible interval
ESS: effective sample size
LBP: low back pain
MCMC: Markov chain Monte Carlo
NH: natural history
NMA: network meta-analysis
NRS: numerical rating scale
PEDro: Physiotherapy Evidence Database scale
PSRF: potential scale reduction factor
SMT: spinal manipulative therapy
VAS: visual analogue scale.

## References

1. Hartvigsen J, Hancock MJ, Kongsted A, Louw Q, Ferreira ML, Genevay S, et al. What low back pain is and why we need to pay attention. Lancet. 2018;391(10137):2356–67.

2. Dieleman JL, Cao J, Chapin A, Chen C, Li Z, Liu A, et al. US Health Care Spending by Payer and Health Condition, 1996-2016. JAMA. 2020;323(9):863–84.

3. Bronfort G, Haas M, Evans R, Leiniger B, Triano J. Effectiveness of manual therapies: the UK evidence report. Chiropr Osteopat. 2010;18(1):3.

4. Cheng M, Xue Y, Cui M, Zeng X, Yang C, Ding F, et al. Global, Regional, and National Burden of Low Back Pain: Findings from the Global Burden of Disease Study 2021 and Projections to 2050. Spine (Phila Pa 1976). 2025.

5. Goetzel RZ, Long SR, Ozminkowski RJ, Hawkins K, Wang S, Lynch W. Health, absence, disability, and presenteeism cost estimates of certain physical and mental health conditions affecting U.S. employers. J Occup Environ Med. 2004;46(4):398–412.

6. Maher C, Underwood M, Buchbinder R. Non-specific low back pain. Lancet. 2017;389(10070):736–47.

7. Artus M, van der Windt DA, Jordan KP, Hay EM. Low back pain symptoms show a similar pattern of improvement following a wide range of primary care treatments: a systematic review of randomized clinical trials. Rheumatology (Oxford). 2010;49(12):2346–56.

8. Barnett AG, van der Pols JC, Dobson AJ. Regression to the mean: what it is and how to deal with it. Int J Epidemiol. 2005;34(1):215–20.

9. Hrobjartsson A, Gotzsche PC. Is the placebo powerless? An analysis of clinical trials comparing placebo with no treatment. N Engl J Med. 2001;344(21):1594–602.

10. Jadhav AB, Jagtap P, Gurav S, Jadhav S, Jadhav N, Akkalkot A. A Survey on Text Mining -Techniques, Application. International Journal of Scientific Research in Computer Science Engineering and Information Technology. 2023:338–43.

11. Inose H, Kato T, Sasaki M, Matsukura Y, Hirai T, Yoshii T, et al. Comparison of decompression, decompression plus fusion, and decompression plus stabilization: a long-term follow-up of a prospective, randomized study. Spine J. 2022;22(5):747–55.

12. Daoust R, Paquet J, Cournoyer A, Piette É, Morris J, Lessard J, et al. Relationship Between Acute Pain Trajectories After an Emergency Department Visit and Chronic Pain: A Canadian Prospective Cohort Study. BMJ Open. 2020;10(12):e040390.

13. Gatchel R, Rosen R, Polatin P. Transitioning from acute to chronic pain: an examination of different trajectories of low-back pain. Healthcare. 2018;6(2):48.

14. Chen Y, Campbell P, Strauss VY, Foster NE, Jordan KP, Dunn KM. Trajectories and predictors of the long-term course of low back pain: cohort study with 5-year follow-up. Pain. 2018;159(2):252–60.

15. Ailliet L, Rubinstein SM, Hoekstra T, van Tulder MW, de Vet HCW. Long-term trajectories of patients with neck pain and low back pain presenting to chiropractic care: A latent class growth analysis. Eur J Pain. 2018;22(1):103–13.

16. Kongsted A, Kent P, Axen I, Downie AS, Dunn KM. What have we learned from ten years of trajectory research in low back pain? BMC Musculoskelet Disord. 2016;17:220.

17. Kongsted A, Andersen CH, Hansen MM, Hestbaek L. Prediction of outcome in patients with low back pain--A prospective cohort study comparing clinicians’ predictions with those of the Start Back Tool. Man Ther. 2016;21:120–7.

18. Downie A, Hancock MJ, Rzewuska M, Williams C, Lin CWC, Maher CG. Trajectories of Acute Low Back Pain. Pain. 2016;157(1):225–34.

19. Meucci RD, Fassa AG, Faria NMX. Prevalence of Chronic Low Back Pain: Systematic Review. Revista De Saúde Pública. 2015;49(0).

20. Gonzalez-Hernandez G, Tahsin T, Goodale BC, Greene AC, Greene CS. Recent Advances and Emerging Applications in Text and Data Mining for Biomedical Discovery. Briefings in Bioinformatics. 2015;17(1):33–42.

21. Dami A, Fakir M, Bouikhalene B. Information Retrieval (IR) and Extracting Associative Rules. Journal of Information Technology Research. 2014;7(4):42–62.

22. Itz CJ, Geurts JW, van Kleef M, Nelemans P. Clinical course of non-specific low back pain: a systematic review of prospective cohort studies set in primary care. Eur J Pain. 2013;17(1):5–15.

23. Hoffmann T, Mar CD, Strong J, Mai J. Patients’ Expectations of Acute Low Back Pain Management: Implications for Evidence Uptake. BMC Family Practice. 2013;14(1).

24. Dunn KM, Campbell P, Jordan KP. Long-term trajectories of back pain: cohort study with 7-year follow-up. BMJ Open. 2013;3(12):e003838.

25. Darlow B, Dowell A, Baxter D, Mathieson F, Perry M, Dean S. The Enduring Impact of What Clinicians Say to People With Low Back Pain. The Annals of Family Medicine. 2013;11(6):527–34.

26. Chapman CR, Fosnocht D, Donaldson GW. Resolution of acute pain following discharge from the emergency department: the acute pain trajectory. J Pain. 2012;13(3):235–41.

27. Axen I, Bodin L, Bergstrom G, Halasz L, Lange F, Lovgren PW, et al. The use of weekly text messaging over 6 months was a feasible method for monitoring the clinical course of low back pain in patients seeking chiropractic care. J Clin Epidemiol. 2012;65(4):454–61.

28. Axen I, Bodin L, Bergstrom G, Halasz L, Lange F, Lovgren PW, et al. Clustering patients on the basis of their individual course of low back pain over a six month period. BMC Musculoskelet Disord. 2011;12:99.

29. Menezes Costa Lda C, Maher CG, McAuley JH, Costa LO. Systematic review of cross-cultural adaptations of McGill Pain Questionnaire reveals a paucity of clinimetric testing. J Clin Epidemiol. 2009;62(9):934–43.

30. Hestbaek L, Kongsted A, Jensen TS, Leboeuf-Yde C. The clinical aspects of the acute facet syndrome: results from a structured discussion among European chiropractors. Chiropr Osteopat. 2009;17:2.

31. Edwards RR. The association of perceived discrimination with low back pain. J Behav Med. 2008;31(5):379–89.

32. Chen C, Hogg-Johnson S, Smith P. The recovery patterns of back pain among workers with compensated occupational back injuries. Occup Environ Med. 2007;64(8):534–40.

33. Shen FH, Samartzis D, Andersson GB. Nonsurgical management of acute and chronic low back pain. J Am Acad Orthop Surg. 2006;14(8):477–87.

34. Crombez G, Vlaeyen JW, Heuts PH, Lysens R. Pain-related fear is more disabling than pain itself: evidence on the role of pain-related fear in chronic back pain disability. Pain. 1999;80(1-2):329–39.

35. Crombez G, Eccleston C, Baeyens F, van Houdenhove B, van den Broeck A. Attention to chronic pain is dependent upon pain-related fear. J Psychosom Res. 1999;47(5):403–10.

36. Epping-Jordan JE, Wahlgren DR, Williams R, Pruitt SD, Slater MA, Patterson TL, et al. Transition to Chronic Pain in Men With Low Back Pain: Predictive Relationships Among Pain Intensity, Disability, and Depressive Symptoms. Health Psychology. 1998;17(5):421–7.

37. Croft PR, Macfarlane GJ, Papageorgiou AC, Thomas E, Silman AJ. Outcome of low back pain in general practice: a prospective study. BMJ. 1998;316(7141):1356–9.

38. Menke JM, Figueredo AJ, Peñaherrera-Aguirre M. The Natural History Trajectory of Acute and Chronic Low Back Pain: A Bayesian Meta-Analysis. Manuscript submitted for publication; 2026. 2026.

39. Kinalski R, Kuwik W, D. P. The comparison of the results of manual therapy versus physiotherapy methods used in treatment of patients with low back pain syndromes. Journal of Manual Medicine. 1989;4(2):44–6.

40. Sturion LA, Nowotny AH, Barillec F, Barette G, Santos GK, Teixeira FA, et al. Comparison between high-velocity low-amplitude manipulation and muscle energy technique on pain and trunk neuromuscular postural control in male workers with chronic low back pain: A randomised crossover trial. S Afr J Physiother. 2020;76(1):1420.

41. Licciardone JC, Gatchel RJ. Osteopathic Medical Care With and Without Osteopathic Manipulative Treatment in Patients With Chronic Low Back Pain: A Pain Registry-Based Study. J Am Osteopath Assoc. 2020;120(2):64–73.

42. Bronfort G, Meier EN, Leininger B, Schneider M, Evans R, Greco C, et al. Spinal Manipulation and Clinician-Supported Biopsychosocial Self-Management for Acute Back Pain: The PACBACK Randomized Clinical Trial. JAMA. 2026;335(6):497–510.

43. Fritz JM, Sharpe J, Greene T, Lane E, Hadizadeh M, McFadden M, et al. Optimization of Spinal Manipulative Therapy Protocols: A Factorial Randomized Trial Within a Multiphase Optimization Framework. J Pain. 2021;22(6):655–68.

44. Whedon JM, Bezdjian S, Dennis P, Fischer VA, Russell R. Cost comparison of two approaches to chiropractic care for patients with acute and sub-acute low Back pain care episodes: a cohort study. Chiropractic & manual therapies. 2020;28(1):68.

45. Goertz CM, Long CR, Vining RD, Pohlman KA, Walter J, Coulter I. Effect of usual medical care plus chiropractic care vs usual medical care alone on pain and disability among US service members with low back pain: a comparative effectiveness clinical trial. JAMA Network Open. 2018;1(1):e180105.

46. Schneider M, Haas M, Glick R, Stevans J, Landsittel D. Comparison of spinal manipulation methods and usual medical care for acute and subacute low back pain: a randomized clinical trial. Spine. 2015;40(4):209–17.

47. Hensel KL, Buchanan S, Brown SK, Rodriguez M, Cruser dA. A multi-site osteopathic healthcare outcomes study assessing the effect of osteopathic manipulative treatment on low back pain in adults: primary outcome results. J Am Osteopath Assoc. 2015;115(3):144–55.

48. Fritz JM, Magel JS, McFadden M, Asche C, Thackeray A, Meier W, et al. Early Physical Therapy vs Usual Care in Patients With Recent-Onset Low Back Pain: A Randomized Clinical Trial. JAMA. 2015;314(14):1459–67.

49. Leemann S, Peterson CK, Schmid C, Anklin B, Humphreys BK. Outcomes of acute and chronic patients with magnetic resonance imaging-confirmed symptomatic lumbar disc herniations receiving high-velocity, low-amplitude, spinal manipulative therapy: a prospective observational cohort study with one-year follow-up. J Manipulative Physiol Ther. 2014;37(3):155–63.

50. von Heymann WJ, Schloemer P, Timm J, Muehlbauer B. Spinal high-velocity low amplitude manipulation in acute nonspecific low back pain: a double-blinded randomized controlled trial in comparison with diclofenac and placebo. Spine (Phila Pa 1976). 2013;38(7):540–8.

51. Goertz CM, Long CR, Hondras MA, Petri R, Delgado R, Lawrence DJ, et al. Adding chiropractic manipulative therapy to standard medical care for patients with acute low back pain: results of a pragmatic randomized comparative effectiveness study. Spine (Phila Pa 1976). 2013;38(8):627–34.

52. Cruser DA, Maurer D, Hensel K, Brown SK, White K, Stoll ST. A randomized, controlled trial of osteopathic manipulative treatment for acute low back pain in active duty military personnel. The Journal of Manual & Manipulative Therapy. 2012;20(1):5–15.

53. Grunnesjö MI, Bogefeldt JP, Blomberg SI, Strender LE, Svärdsudd KF. A randomized controlled trial of the effects of muscle stretching, manual therapy and steroid injections in addition to ’stay active’ care on health-related quality of life in acute or subacute low back pain. Clin Rehabil. 2011;25(11):999–1010.

54. Juni P, Battaglia M, Nuesch E, Hammerle G, Eser P, van Beers R, et al. A randomised controlled trial of spinal manipulative therapy in acute low back pain. Ann Rheum Dis. 2009;68(9):1420–7.

55. Hallegraeff JM, de Greef M, Winters JC, Lucas C. Manipulative therapy and clinical prediction criteria in treatment of acute nonspecific low back pain. Percept Mot Skills. 2009;108(1):196–208.

56. Cleland JA, Fritz JM, Kulig K, Davenport TE, Eberhart S, Magel J, et al. Comparison of the effectiveness of three manual physical therapy techniques in a subgroup of patients with low back pain who satisfy a clinical prediction rule: a randomized clinical trial. Spine. 2009;34(25):2720–9.

57. Paatelma M, Kilpikoski S, Simonen R, Heinonen A, Alen M, Videman T. Orthopaedic manual therapy, McKenzie method or advice only for low back pain in working adults: a randomized controlled trial with one year follow-up. Journal of Rehabilitation Medicine. 2008;40(10):858–63.

58. Hancock MJ, Maher CG, Latimer J, McLachlan AJ, Cooper CW, Day RO, et al. Assessment of diclofenac or spinal manipulative therapy, or both, in addition to recommended first-line treatment for acute low back pain: a randomised controlled trial. Lancet. 2007;370(9599):1638–43.

59. Santilli V, Beghi E, Finucci S. Chiropractic manipulation in the treatment of acute back pain and sciatica with disc protrusion: a randomized double-blind clinical trial of active and simulated spinal manipulations. Spine J. 2006;6(2):131–7.

60. Beyerman KL, Palmerino MB, Zohn LE, Kane GM, Foster KA. Efficacy of treating low back pain and dysfunction secondary to osteoarthritis: chiropractic care compared with moist heat alone. J Manipulative Physiol Ther. 2006;29(2):107–14.

61. Hawk C, Long CR, Rowell RM, Gudavalli MR, Jedlicka J. A randomized trial investigating a chiropractic manual placebo: a novel design using standardized forces in the delivery of active and control treatments. J Altern Complement Med. 2005;11(1):109–17.

62. Wand BM, Bird C, McAuley JH, Dore CJ, MacDowell M, De Souza LH. Early intervention for the management of acute low back pain: a single-blind randomized controlled trial of biopsychosocial education, manual therapy, and exercise. Spine. 2004;29(21):2350–6.

63. UK BEAM TT. United Kingdom back pain exercise and manipulation (UK BEAM) randomised trial: effectiveness of physical treatments for back pain in primary care. BMJ. 2004;329(7479):1377–85.

64. Hoiriis KT, Pfleger B, McDuffie FC, Cotsonis G, Elsangak O, Hinson R, et al. A randomized clinical trial comparing chiropractic adjustments to muscle relaxants for subacute low back pain. Journal of Manipulative and Physiological Therapeutics. 2004;27(6):388–98.

65. Grunnesjö MI, Bogefeldt JP, Svärdsudd KF, Blomberg SIE. A randomized controlled clinical trial of stay-active care versus manual therapy in addition to stay-active care: functional variables and pain. Journal of Manipulative and Physiological Therapeutics. 2004;27(7):431–41.

66. Williams NH, Wilkinson C, Russell I, Edwards RT, Hibbs R, Linck P, et al. Randomized osteopathic manipulation study (ROMANS): pragmatic trial for spinal pain in primary care. Family Practice. 2003;20(6):662–9.

67. Hurwitz EL, Morgenstern H, Harber P, Kominski GF, Belin TR, Yu F, et al. A randomized trial of medical care with and without physical therapy and chiropractic care with and without physical modalities for patients with low back pain: 6-month follow-up outcomes from the UCLA low back pain study. Spine (Phila Pa 1976). 2002;27(20):2193–204.

68. Hsieh CY, Adams AH, Tobis J, Hong CZ, Danielson C, Platt K, et al. Effectiveness of four conservative treatments for subacute low back pain: a randomized clinical trial. Spine (Phila Pa 1976). 2002;27(11):1142–8.

69. Morton JE. Manipulation in the treatment of acute low back pain. Journal of Manual & Manipulative Therapy. 1999;7(4):182–9.

70. Andersson GB, Lucente T, Davis AM, Kappler RE, Lipton JA, Leurgans S. A comparison of osteopathic spinal manipulation with standard care for patients with low back pain. N Engl J Med. 1999;341(19):1426–31.

71. Skargren EI, Oberg BE. Predictive factors for 1-year outcome of low-back and neck pain in patients treated in primary care: comparison between the treatment strategies chiropractic and physiotherapy. Pain. 1998;77(2):201–7.

72. Cherkin DC, Deyo RA, Battie M, Street J, Barlow W. A comparison of physical therapy, chiropractic manipulation, and provision of an educational booklet for the treatment of patients with low back pain. N Engl J Med. 1998;339(15):1021–9.

73. Skargren EI, Oberg BE, Carlsson PG, Gade M. Cost and effectiveness analysis of chiropractic and physiotherapy treatment for low back and neck pain. Six-month follow-up. Spine (Phila Pa 1976). 1997;22(18):2167–77.

74. Meade TW, Dyer S, Browne W, Frank AO. Randomised comparison of chiropractic and hospital outpatient management for low back pain: results from extended follow up. BMJ. 1995;311(7001):349–51.

75. Pope MH, Phillips RB, Haugh LD, Hsieh CY, MacDonald L, Haldeman S. A prospective randomized three-week trial of spinal manipulation, transcutaneous muscle stimulation, massage and corset in the treatment of subacute low back pain. Spine (Phila Pa 1976). 1994;19(22):2571–7.

76. Erhard RE, Delitto A, Cibulka MT. Relative effectiveness of an extension program and a combined program of manipulation and flexion and extension exercises in patients with acute low back syndrome. Phys Ther. 1994;74(12):1093–100.

77. Blomberg S, Hallin G, Grann K, Berg E, Sennerby U. Manual therapy with steroid injections--a new approach to treatment of low back pain. A controlled multicenter trial with an evaluation by orthopedic surgeons. Spine. 1994;19(5):569–77.

78. Delitto A, Cibulka MT, Erhard RE, Bowling RW, Tenhula JA. Evidence for use of an extension-mobilization category in acute low back syndrome: a prescriptive validation pilot study. Phys Ther. 1993;73(4):216–22; discussion 23–8.

79. Cramer GD, Humphreys CR, Hondras MA, McGregor M, Triano JJ. The Hmax/Mmax ratio as an outcome measure for acute low back pain. J Manipulative Physiol Ther. 1993;16(1):7– 13.

80. Wreje U, Nordgren B, Aberg H. Treatment of pelvic joint dysfunction in primary care--a controlled study. Scand J Prim Health Care. 1992;10(4):310–5.

81. Hsieh CY, Phillips RB, Adams AH, Pope MH. Functional outcomes of low back pain: comparison of four treatment groups in a randomized controlled trial. J Manipulative Physiol Ther. 1992;15(1):4–9.

82. Blomberg S, Svardsudd K, Mildenberger F. A controlled, multicentre trial of manual therapy in low-back pain. Initial status, sick-leave and pain score during follow-up. Scand J Prim Health Care. 1992;10(3):170–8.

83. Herzog W, Conway PJW, Willcox BJ. Effects of different treatment modalities on gait symmetry and clinical measures for sacroiliac joint patients. J Manipulative Physiol Ther. 1991;14:104–9.

84. MacDonald RS, Bell CM. An open controlled assessment of osteopathic manipulation in nonspecific low-back pain. Acute and subacute. Spine. 1990;15(5):364–70.

85. Postacchini F, Facchini M, Palieri P. Efficacy of various forms of conservative treatment in low back pain: a comparative study. Neuro-Orthopedics. 1988;6:28–35.

86. Mathews JA, Mills SB, Jenkins VM, Grimes SM, Morkel MJ, Mathews W, et al. Back pain and sciatica: controlled trials of manipulation, traction, sclerosant and epidural injections, Trial B. Br J Rheumatol. 1987 b;26(6):416–23.

87. Mathews JA, Mills SB, Jenkins VM, Grimes SM, Morkel MJ, Mathews W, et al. Back pain and sciatica: controlled trials of manipulation, traction, sclerosant and epidural injections, Trial A. Br J Rheumatol. 1987 a;26(6):416–23.

88. Hadler NM, Curtis P, Gillings DB, Stinnett S. A benefit of spinal manipulation as adjunctive therapy for acute low-back pain: a stratified controlled trial. Spine (Phila Pa 1976). 1987;12(7):702–6.

89. Waterworth RF, Hunter IA. An open study of diflunisal, conservative and manipulative therapy in the management of acute mechanical low back pain. NZ Med J. 1985;98(779):372–5.

90. Rupert RL, Ezzeldin MT. Chiropractic adjustments: results of a controlled clinical trial in Egypt. ICA International Review of Chiropractic. 1985;Winter:58–60.

91. Gibson T, Grahame R, Harkness J, Woo P, Blagrave P, Hills R. Controlled comparison of short-wave diathermy treatment with osteopathic treatment in non-specific low back pain. Lancet. 1985;1(8440):1258–61.

92. Godfrey CM, Morgan PP, Schatzker J. A randomized trial of manipulation for low-back pain in a medical setting. Spine (Phila Pa 1976). 1984;9(3):301–4.

93. Farrell JP, Twomey LT. Acute low back pain. Comparison of two conservative treatment approaches. Med J Aust. 1982;1(4):160–4.

94. Hoehler FK, Tobis JS, Buerger AA. Spinal manipulation for low back pain. JAMA. 1981;245(18):1835–8.

95. Coxhead CE, Inskip H, Meade TW, North WR, Troup JD. Multicentre trial of physiotherapy in the management of sciatic symptoms. Lancet. 1981;1(8229):1065–8.

96. Rasmussen GG. Manipulation in treatment of low back pain: a randomized clinical trial. Manuelle Medizin. 1979;1:8–10.

97. Sims-Williams H, Jayson MI, Young SM, Baddeley H, Collins E. Controlled trial of mobilisation and manipulation for patients with low back pain in general practice. Br Med J. 1978;2(6148):1338–40.

98. Evans DP, Burke MS, Lloyd KN, Roberts EE, Roberts GM. Lumbar spinal manipulation on trial. Part I--clinical assessment. Rheumatol Rehabil. 1978;17(1):46–53.

99. Bergquist-Ullman M, Larsson U. Acute low back pain in industry. A controlled prospective study with special reference to therapy and confounding factors. Acta Orthop Scand. 1977(170):1–117.

100. Aboagye E, Hansson E, Hagberg J, Axen I. Cost-effectiveness of early interventions for non-specific low back pain: a randomized controlled trial. Trials. 2022;23(1):669.

101. Alkhathami K, Alshehre Y, Brizzolara K, Weber M, Wang-Price S. Effectiveness of Spinal Stabilization Exercises on Movement Performance in Adults with Chronic Low Back Pain. Int J Sports Phys Ther. 2023;18(1):169–72.

102. Anderson BD, Mayer JM, Graves JE. Changes in muscle activation following spinal manipulation in patients with chronic low back pain: a pilot study. J Manipulative Physiol Ther. 2024;47(1):12–20.

103. Auger C, Demers M, Sweeney A. The effect of adding osteopathic treatment to standard care for patients with chronic low back pain: a pilot randomized controlled trial. Int J Osteopath Med. 2020;38:24–33.

104. Aure OF, Nilsen JH, Vasseljen O. Manual therapy and exercise therapy in patients with chronic low back pain: a randomized, controlled trial with 1-year follow-up. Spine (Phila Pa 1976). 2003;28(6):525–31; discussion 31–2.

105. Balthazard P, de Goumoens P, Rivier G, Demeulenaere P, Ballabeni P, Deriaz O. Manual therapy followed by specific active exercises versus a placebo followed by specific active exercises on the improvement of functional disability in patients with chronic non specific low back pain: a randomized controlled trial. BMC Musculoskelet Disord. 2012;13:162.

106. Bicalho E, Setti JA, Macagnan J, Cano JL, Manffra EF. Immediate effects of a high-velocity spine manipulation in paraspinal muscles activity of nonspecific chronic low-back pain subjects. Man Ther. 2010;15(5):469–75.

107. Blanco Giménez V, Jiménez Rejano JJ, Chillón Martínez R. Effects of thrust joint manipulation versus non-thrust manual therapy in patients with chronic low back pain: a randomized clinical trial. Arch Phys Med Rehabil. 2024;105(7):1248–56.

108. Boff TA, Pasinato F, Ben AJ, Bosmans JE, van Tulder M, Carregaro RL. Effectiveness of spinal manipulation and myofascial release compared with spinal manipulation alone on health-related outcomes in individuals with non-specific low back pain: randomized controlled trial. Physiotherapy. 2020;107:71–80.

109. Bronfort G, Goldsmith CH, Nelson CF, Boline PD, Anderson AV. Trunk exercise combined with spinal manipulative or NSAID therapy for chronic low back pain: a randomized, observer-blinded clinical trial. J Manipulative Physiol Ther. 1996;19(9):570–82.

110. Burton AK, Tillotson KM, Cleary J. Single-blind randomised controlled trial of chemonucleolysis and manipulation in the treatment of symptomatic lumbar disc herniation. Eur Spine J. 2000;9(3):202–7.

111. Cecchi F, Molino-Lova R, Chiti M, Pasquini G, Paperini A, Conti AA, et al. Spinal manipulation compared with back school and with individually delivered physiotherapy for the treatment of chronic low back pain: a randomized trial with one-year follow-up. Clinical Rehabilitation. 2010;24(1):26–36.

112. Chiradejnant A, Maher CG, Latimer J, Stepkovitch N. Efficacy of “therapist-selected” versus “randomly selected” mobilisation techniques for the treatment of low back pain: a randomised controlled trial. Aust J Physiother. 2003;49(4):233–41.

113. Chow RK, Ng GY, Cheng SW. Effectiveness of multidisciplinary treatment for patients with chronic low back pain: a randomised clinical trial. Hong Kong Physiother J. 2021;41(1):15–25.

114. Chown M, Whittamore L, Rush M, Allan S, Stott D, Archer M. A prospective study of patients with chronic back pain randomised to group exercise, physiotherapy or osteopathy. Physiotherapy. 2008;94(1):21–8.

115. de Oliveira Meirelles F, de Oliveira Muniz Cunha JC, da Silva EB. Osteopathic manipulation treatment versus therapeutic exercises in patients with chronic nonspecific low back pain: A randomized, controlled and double-blind study. J Back Musculoskelet Rehabil. 2020;33(3):367–77.

116. Donaldson M, Learman K, O’Halloran B, Showalter C, Cook C. The role of patients’ expectation of appropriate initial manual therapy treatment in outcomes for patients with low back pain. Journal of Manipulative and Physiological Therapeutics. 2016;39(4):276–83.

117. Doran DM, Newell DJ. Manipulation in treatment of low back pain: a multicentre study. Br Med J. 1975;2(5964):161–4.

118. Ferreira ML, Ferreira PH, Latimer J, Herbert RD, Hodges PW, Jennings MD, et al. Comparison of general exercise, motor control exercise and spinal manipulative therapy for chronic low back pain: A randomized trial. Pain. 2007;131(1-2):31–7.

119. Geisser ME, Wiggert EA, Haig AJ, Colwell MO. A randomized, controlled trial of manual therapy and specific adjuvant exercise for chronic low back pain. The Clinical Journal of Pain. 2005;21(6):463–70.

120. Gevers-Montoro C, Romero-Santiago B, Medina-Garcia I, Larranaga-Arzamendi B, Alvarez-Galovich L, Ortega-De Mues A, et al. Reduction of Chronic Primary Low Back Pain by Spinal Manipulative Therapy is Accompanied by Decreases in Segmental Mechanical Hyperalgesia and Pain Catastrophizing: A Randomized Placebo-controlled Dual-blind Mixed Experimental Trial. J Pain. 2024;25(8):104500.

121. Ghroubi S, Elleuch H, Baklouti S, Elleuch MH. Chronic low back pain and vertebral manipulation. Annales de Réadaptation et de Médecine Physique. 2007;50(7):570–6.

122. Giles LG, Muller R. Chronic spinal pain syndromes: a clinical pilot trial comparing acupuncture, a nonsteroidal anti-inflammatory drug, and spinal manipulation. J Manipulative Physiol Ther. 1999;22(6):376–81.

123. Giles LG, Muller R. Chronic spinal pain: a randomized clinical trial comparing medication, acupuncture, and spinal manipulation. Spine (Phila Pa 1976). 2003;28(14):1490–502; discussion 502–3.

124. Glover JR, Morris JG, Khosla T. Back pain: a randomized clinical trial of rotational manipulation of the trunk. Br J Ind Med. 1974;31(1):59–64.

125. Goldby LJ, Moore AP, Doust J, Trew ME. A randomized controlled trial investigating the efficiency of musculoskeletal physiotherapy on chronic low back disorder. Spine. 2006;31(10):1083–93.

126. Gudavalli MR, Cambron JA, McGregor M, Jedlicka J, Keenum M, Ghanayem AJ, et al. A randomized clinical trial and subgroup analysis to compare flexion-distraction with active exercise for chronic low back pain. Eur Spine J. 2006;15(7):1070–82.

127. Haas M, Vavrek D, Peterson D, Polissar N, Neradilek MB. Dose-response and efficacy of spinal manipulation for care of chronic low back pain: a randomized controlled trial. The Spine Journal. 2014;14(7):1106–16.

128. Hadizadeh M, Jafarnezhadgero AA, Lotfian S, Seyed Majidi MH. The impacts of cognitive functional therapy on pain, lumbar kinematics and trunk muscle activity in chronic non-specific low back pain during walking: a randomized controlled trial. Musculoskelet Sci Pract. 2020;46:102109.

129. Hartz CS, Almeida de Molon Mendes M, Buck KH, Moreno MA, Bortolazzo GL. Effect of osteopathic manipulative treatment on pain, flexibility, autonomic modulation of heart rate, energy and thermal profile in patients with chronic low back pain. blind randomized clinical trial. J Bodyw Mov Ther. 2025;44:452–61.

130. Hemmila HM, Keinanen-Kiukaanniemi SM, Levoska S, Puska P. Does folk medicine work? A randomized clinical trial on patients with prolonged back pain. Archives of Physical Medicine and Rehabilitation. 1997;78(6):571–7.

131. Hemmila HM, Keinanen-Kiukaanniemi SM, Levoska S, Puska P. Long-term effectiveness of bone-setting, light exercise therapy, and physiotherapy for prolonged back pain: a randomized controlled trial. J Manipulative Physiol Ther. 2002;25(2):99–104.

132. Hondras MA, Long CR, Cao Y, Rowell RM, Meeker WC. A randomized controlled trial comparing 2 types of spinal manipulation and minimal conservative medical care for adults 55 years and older with subacute or chronic low back pain. Journal of Manipulative and Physiological Therapeutics. 2009;32(5):330–43.

133. Koes BW, Bouter LM, van Mameren H. Randomised clinical trial of manipulative therapy and physiotherapy for persistent back and neck complaints: results of one year follow up. BMJ. 1992b;304(6827):601–5.

134. Koes BW, Bouter LM, van Mameren H, Essers AH, Verstegen GM, Hofhuizen DM, et al. The effectiveness of manual therapy, physiotherapy, and treatment by the general practitioner for nonspecific back and neck complaints. A randomized clinical trial. Spine. 1992a;17(1):28–35.

135. Krekoukias G, Gelalis ID, Xenakis T, Gioftsos G, Dimitriadis Z, Sakellari V. Spinal mobilization vs conventional physiotherapy in the management of chronic low back pain due to spinal disk degeneration: a randomized controlled trial. Journal of Manual & Manipulative Therapy. 2017;25(2):66–73.

136. Licciardone JC, Minotti DE, Gatchel RJ, Kearns CM, Singh KP. Osteopathic manual treatment and ultrasound therapy for chronic low back pain: a randomized controlled trial. Ann Fam Med. 2013;11(2):122–9.

137. Licciardone JC, Stoll ST, Fulda KG, Russo DP, Siu J, Winn W, et al. Osteopathic manipulative treatment for chronic low back pain: a randomized controlled trial. Spine (Phila Pa 1976). 2003;28(13):1355–62.

138. Mandara A, Fusaro A, Musicco M, Bado F. A randomised controlled trial on the effectiveness of osteopathic manipulative treatment of chronic low back pain. International Journal of Osteopathic Medicine. 2008;11:149–68.

139. McCarthy CJ, Potter L, Oldham JA. Comparing targeted thrust manipulation with general thrust manipulation in patients with low back pain. A general approach is as effective as a specific one. A randomised controlled trial. BMJ Open Sport Exerc Med. 2019;5(1):e000514.

140. Mohseni-Bandpei MA, Critchley J, Staunton T, Richardson B. A prospective randomised controlled trial of spinal manipulation and ultrasound in the treatment of chronic low back pain. Physiotherapy. 2006;92(1):34–42.

141. Muller R, Giles LGF. Long-term follow-up of a randomized clinical trial assessing the efficacy of medication, acupuncture, and spinal manipulation for chronic mechanical spinal pain syndromes. Journal of Manipulative and Physiological Therapeutics. 2005;28(1):3–11.

142. Nguyen C, Boutron I, Zegarra-Parodi R, Baron G, Alami S, Sanchez K, et al. Effect of Osteopathic Manipulative Treatment vs Sham Treatment on Activity Limitations in Patients With Nonspecific Subacute and Chronic Low Back Pain: A Randomized Clinical Trial. JAMA Intern Med. 2021;181(5):620–30.

143. Nguyen C, Palazzo C, Grabar S. Osteopathic manipulative treatment combined with conventional care versus conventional care alone for patients with chronic nonspecific low back pain: a multicenter randomized controlled trial. Ther Adv Chronic Dis. 2021;12:20406223211011017.

144. Niemisto L, Lahtinen-Suopanki T, Rissanen P, Lindgren KA, Sarna S, Hurri H. A randomized trial of combined manipulation, stabilizing exercises, and physician consultation compared to physician consultation alone for chronic low back pain. Spine (Phila Pa 1976). 2003;28(19):2185–91.

145. Ongley MJ, Klein RG, Dorman TA, Eek BC, Hubert LJ. A new approach to the treatment of chronic low back pain. Lancet. 1987;2(8551):143–6.

146. Rasmussen-Barr E, Nilsson-Wikmar L, Arvidsson I. Stabilizing training compared with manual treatment in sub-acute and chronic low-back pain. Man Ther. 2003;8(4):233–41.

147. Ritvanen T, Zaproudina N, Nissen M, Leinonen V, Hänninen O. Dynamic surface electromyographic responses in chronic low back pain treated by traditional bone setting and conventional physical therapy. Journal of Manipulative and Physiological Therapeutics. 2007;30(1):31–7.

148. Schulz C, Evans R, Maiers M, Schulz K, Leininger B, Bronfort G. Spinal manipulative therapy and exercise for older adults with chronic low back pain: a randomized clinical trial. Chiropractic & manual therapies. 2019;27:21.

149. Senna MK, Machaly SA. Does maintained spinal manipulation therapy for chronic nonspecific low back pain result in better long-term outcome? Spine (Phila Pa 1976). 2011;36(18):1427–37.

150. Shirado O, Doi T, Akai M, Hoshino Y, Fujino K, Hayashi K, et al. Multicenter randomized controlled trial to evaluate the effect of home-based exercise on patients with chronic low back pain: the Japan low back pain exercise therapy study. Spine (Phila Pa 1976). 2010;35(17):E811–9.

151. Sims-Williams H, Jayson MI, Young SM, Baddeley H, Collins E. Controlled trial of mobilisation and manipulation for low back pain: hospital patients. Br Med J. 1979;2(6201):1318–20.

152. Skillgate E, Vingård E, Alfredsson L. Naprapathic manual therapy or evidence-based care for back and neck pain: a randomized, controlled trial. The Clinical Journal of Pain. 2007;23(5):431–9.

153. Tavares FAG, Rossiter JVA, Lima GCL, de Oliveira LG, Cavalcante WS, Avila MA, et al. Additional effect of pain neuroscience education to spinal manipulative therapy on pain and disability for patients with chronic low back pain: a randomized controlled trial. Braz J Phys Ther. 2023;27(5):100555.

154. Teodorczyk-Injeyan JA, Triano JJ, Gringmuth R, DeGraauw C, Chow A, Injeyan HS. Effects of spinal manipulative therapy on inflammatory mediators in patients with non-specific low back pain: a non-randomized controlled clinical trial. Chiropractic & manual therapies. 2021;29(1):3.

155. Thomas JS, Clark BC, Russ DW, France CR, Ploutz-Snyder R, Corcos DM, et al. Effect of Spinal Manipulative and Mobilization Therapies in Young Adults With Mild to Moderate Chronic Low Back Pain: A Randomized Clinical Trial. JAMA Netw Open. 2020;3(8):e2012589.

156. Timm KE. A randomized-control study of active and passive treatments for chronic low back pain following L5 laminectomy. J Orthop Sports Phys Ther. 1994;20(6):276–86.

157. Triano JJ, McGregor M, Hondras MA, Brennan PC. Manipulative therapy versus education programs in chronic low back pain. Spine (Phila Pa 1976). 1995;20(8):948–55.

158. Vismara L, Cimolin V, Menegoni F, Zaina F, Galli M, Negrini S, et al. Osteopathic manipulative treatment in obese patients with chronic low back pain: a pilot study. Man Ther. 2012;17(5):451–5.

159. Waagen GN, Haldeman S, Cook G, Lopez D, DeBoer KF. Short term trial of chiropractic adjustments for the relief of chronic low back pain. Manual Medicine. 1986;2:63–7.

160. Waqqar S, Shakil-ur-Rehman S, Ahmad S. McKenzie treatment versus mulligan sustained natural apophyseal glides for chronic mechanical low back pain. Pakistan Journal of Medical Sciences. 2016;32(2):476–9.

161. Wilkey A, Gregory M, Byfield D, McCarthy PW. A comparison between chiropractic management and pain clinic management for chronic low-back pain in a national health service outpatient clinic. J Altern Complement Med. 2008;14(5):465–73.

162. Zaproudina N, Hietikko T, Hänninen OOP, Airaksinen O. Effectiveness of traditional bone setting in treating chronic low back pain: a randomised pilot trial. Complementary Therapies in Medicine. 2009;17(1):23–8.

163. Zylbergold RS, Piper MC. Lumbar disc disease: comparative analysis of physical therapy treatments. Arch Phys Med Rehabil. 1981;62(4):176–9.

164. Deyo RA, Battie M, Beurskens AJ, Bombardier C, Croft P, Koes B, et al. Outcome measures for low back pain research. A proposal for standardized use. Spine (Phila Pa 1976). 1998;23(18):2003–13.

165. Hedges LV. Distribution theory for Glass’s estimator of effect size and related estimators. Journal of Educational Statistics. 1981;6(2):107–28.

166. Furukawa TA, Cipriani A, Barbui C, Brambilla P, Watanabe N. Imputing response rates from means and standard deviations in meta-analyses. International Clinical Psychopharmacology. 2005;20(1):49–52.

167. Anzures-Cabrera J, Sarpatwari A, Higgins JPT. Expressing findings from meta-analyses of continuous outcomes in terms of risks. Statistics in Medicine. 2011;30(25):2967–85.

168. Ostelo RWJG, Deyo RA, Stratford P, Waddell G, Croft P, Von Korff M, et al. Interpreting change scores for pain and functional status in low back pain: towards international consensus regarding minimal important change. Spine. 2008;33(1):90–4.

169. Briggs A, Claxton K, Sculpher M. Decision Modelling for Health Economic Evaluation. Oxford: Oxford University Press; 2006.

170. Kruschke JK. Rejecting or accepting parameter values in Bayesian estimation. Advances in Methods and Practices in Psychological Science. 2018;1(2):270–80.

171. Salanti G, Schmid CH. Research Synthesis Methods special issue on network meta-analysis: introduction from the editors. Res Synth Methods. 2012;3(2):69–70.

172. Dias S, Ades AE, Welton NJ, Jansen JP, Sutton AJ. Network Meta-Analysis for Decision-Making. Hoboken, NJ: Wiley; 2018.

173. Higgins JP, Jackson D, Barrett JK, Lu G, Ades AE, White IR. Consistency and inconsistency in network meta-analysis: concepts and models for multi-arm studies. Res Synth Methods. 2012;3(2):98–110.

174. Dias S, Welton NJ, Caldwell DM, Ades AE. Checking consistency in mixed treatment comparison meta-analysis. Statistics in Medicine. 2010;29(7-8):932–44.

175. Kriegeskorte N, Simmons WK, Bellgowan PSF, Baker CI. Circular analysis in systems neuroscience: the dangers of double dipping. Nature Neuroscience. 2009;12(5):535–40.

176. Plummer M, editor JAGS: A program for analysis of Bayesian graphical models using Gibbs sampling. Proceedings of the 3rd International Workshop on Distributed Statistical Computing (DSC 2003); 2003; Vienna, Austria.

177. Gelman A, Rubin DB. Inference from iterative simulation using multiple sequences. Statistical Science. 1992;7(4):457–72.

178. Brooks SP, Gelman A. General methods for monitoring convergence of iterative simulations. Journal of Computational and Graphical Statistics. 1998;7(4):434–55.

179. Salanti G. Indirect and mixed-treatment comparison, network, or multiple-treatments meta-analysis: many names, many benefits, many concerns for the next generation evidence synthesis tool. Res Synth Methods. 2012;3(2):80–97.

180. Rothwell PM. External validity of randomised controlled trials: “to whom do the results of this trial apply?”. Lancet. 2005;365(9453):82–93.

181. Gelman A, Carlin JB, Stern HS, Dunson DB, Vehtari A, Rubin DB. Bayesian Data Analysis, Third Edition. 3rd ed. Boca Raton, FL: CRC Press; 2013.

182. Menke JM. Do manual therapies help low back pain? A comparative effectiveness meta-analysis. Spine. 2014;39(7):E463–E72.

183. Ernst E, Resch KL. Concept of true and perceived placebo effects. BMJ. 1995;311(7004):551–3.

184. Humphrey N. Great expectations: The evolutionary psychology of faith healing and the placebo effect. In: C. von Hofsten C, Bäckman L, editors. Psychology at the Turn of the Millennium, Volume 2: Social, developmental, and clinical perspectives (pp 225–246) 2. New York, NY: Taylor & Frances/Routledge; 2002. p. 225–46.

185. Trimmer PC, Marshall JAR, Fromhage L, McNamara JM, Houston AI. Understanding the placebo effect from an evolutionary perspective. Evol Hum Behav. 2013;34(1):8–15.

186. Koban L, Jepma M, Geuter S, Wager TD. What’s in a Word? How Instructions, Suggestions, and Social Information Change Pain and Emotion. Neuroscience & Biobehavioral Reviews. 2017;81:29–42.

187. Hohenschurz-Schmidt D, Draper-Rodi J, Vase L, Scott W, McGregor A, Soliman N, et al. Blinding and sham control methods in trials of physical, psychological, and self-management interventions for pain (article I): a systematic review and description of methods. Pain. 2023;164(3):469–84.

188. Trinquart L, Attiche N, Bafeta A, Porcher R, Ravaud P. Uncertainty in treatment rankings: reanalysis of network meta-analyses of randomized trials. Annals of internal medicine. 2016;164(10):666–73.

189. Riley RD, Higgins JPT, Deeks JJ. Interpretation of random effects meta-analyses. BMJ. 2011;342:d549.

190. Altman DG, Bland JM. Absence of evidence is not evidence of absence. BMJ. 1995;311(7003):485.

191. Henschke N, Maher CG, Refshauge KM, Herbert RD, Cumming RG, Bleasel J, et al. Prognosis in patients with recent onset low back pain in Australian primary care: inception cohort study. BMJ. 2008;337(7662):a171.

192. Costa Lda C, Maher CG, McAuley JH, Hancock MJ, Herbert RD, Refshauge KM, et al. Prognosis for patients with chronic low back pain: inception cohort study. BMJ. 2009;339:b3829.

193. Probst JC, Laditka SB, Wang JY, Johnson AO. Effects of residence and race on burden of travel for care: cross sectional analysis of the 2001 US National Household Travel Survey. BMC Health Serv Res. 2007;7:40.

194. Ray KN, Chari AV, Engberg J, Bertolet M, Mehrotra A. Opportunity costs of ambulatory medical care in the United States. Am J Manag Care. 2015;21(8):567–74.

195. Service IR. Announcement 2026-11: revised optional standard mileage rates for 2026. Internal Revenue Bulletin [Internet]. 2026 October 22, 2026. Available from: https://www.irs.gov/pub/irs-irbs/irb26-29.pdf.

196. Parry IWH, Walls M, Harrington W. Automobile externalities and policies. Journal of Economic Literature. 2007;45(2):373–99.

197. Chen B, Brown HS, Douphrate D, Janak J, Gabriel KP, Peng T. Predictors of Utilization Frequency of and Expenditure Amount for Chiropractic Care in U.S. Adults: A Retrospective Cohort Study. J Manipulative Physiol Ther. 2025;48(1-5):10–26.

198. Ernst E. Adverse effects of spinal manipulation: a systematic review. J R Soc Med. 2007;100(7):330–8.

199. Hebert JJ, Stomski NJ, French SD, Rubinstein SM. Serious Adverse Events and Spinal Manipulative Therapy of the Low Back Region: A Systematic Review of Cases. J Manipulative Physiol Ther. 2015;38(9):677–91.

200. Chen B, Brown HS, Douphrate D, Janak J, Gabriel KP, Peng T. National Trends in the Utilization and Expenditure of Chiropractic Care in U.S. Adults: Analysis of the 2007-2016 Medical Expenditure Panel Survey. J Manipulative Physiol Ther. 2025;48(1-5):1–9.

201. Jenks A, de Zoete A, van Tulder M, Rubinstein SM, International IPDSMTg. Spinal manipulative therapy in older adults with chronic low back pain: an individual participant data meta-analysis. Eur Spine J. 2022;31(7):1821–45.

202. de Zoete A, Rubinstein SM, de Boer MR. The effect of spinal manipulative therapy on pain relief and function in patients with chronic low back pain: an individual participant data meta-analysis. Physiotherapy. 2021;111:58–65.

203. Servick K. Outsmarting the placebo effect. Science. 2014;345(6203):1446–7.

